# Covid-19 and the South Asian Countries: factors ruling the pandemic

**DOI:** 10.1101/2021.05.04.21256590

**Authors:** Tannishtha Biswas, Madhura Mondal, Srijan Bhattacharya, Moitrayee Sarkar, Bikram Dhara, Arup Kumar Mitra, Ayan Chandra

## Abstract

The novel corona virus causing Covid-19 was first detected in the city of Wuhan, China in December, 2019. In matter of months Covid-19 was declared a pandemic by the World Health Organization. The focus of this research includes the probable factors that might have played an important role in the spread of this infection causing a global threat. In this study we dealt with the South Asian countries namely Bangladesh, Bhutan, India, Maldives, Nepal, Pakistan. Data on the demography of the countries, the climatic and geographical conditions, the socio-economic statuses, GDP being in the forefront, was collected and compared with Covid-19 related data such as total number of positive, recovered and death cases, etc. to determine if there was any significant correlation. The wide range of correlations observed can curve the path for the future research to understand the factors behind the spread of the communicable disease, analyzing the dynamics of the future biological threats to mankind and design the precautionary or preventive methods accordingly.

## Introduction

The 2019 novel coronavirus (2019-nCoV) or the severe acute respiratory syndrome coronavirus 2 (SARS-CoV-2) has been spreading at a threatening rate giving the world a new health crisis to deal with. The transmission of the virus occurs when infected droplets are inhaled or contacted. It takes 2 to 14 days as incubation period and to display symptoms. The symptoms include fever, dry cough, sore throat, breathlessness, fatigue, malaise. China shows a fatality rate of 2-3% however the disease is spreading heavily in South Asian countries, giving nightmares on overwhelming health systems. The first country to report a confirmed case in South Asia was Nepal which documented its first case on 23^rd^ January 2020, in a man who had returned from China on 5 January (Fauci *et.al*., 2020). in India, the first case of COVID-19 was reported on 30 January, 2020. As of 15 June 2020, the Ministry of Health and Family Welfare (MoHFW) has confirmed a total of 332,424 cases, 169,798 recoveries (including 1 migration) and 9,520 deaths in the country. India currently has the largest number of confirmed cases in Asia, and has the fourth highest number of confirmed cases in the world with the number of total confirmed cases crossed the 100,000 and 200,000 marks on 19^th^ May and 3^rd^ June, 2020, respectively. India’s case fatality rate is relatively lower at 2.80%, against the global 6.13%, as of 3 June. On June 10, India’s recoveries exceeded active cases for the first time reducing 49% of total infections. The other South Asian countries Bangladesh, Bhutan, Maldives, Pakistan and Sri Lanka reported their first Covid cases on 8^th^ March 2020, 6^th^ March2020, 7^th^ March 2020, 26^th^ February 2020, and 27^th^ January 2020 respectively. Thereafter with the rise in the number of cases, lockdown was imposed, to curb community transmission, on the following countries on the following dates: India – 25^th^ March 2020, Pakistan – 1^st^ April 2020, Bangladesh – 26^th^ March 2020, and Nepal – 24^th^ March, 2020.^[2]^ The effectiveness of the lockdown in ‘flattening the curve’ was also procured. A comparative study of the testing rate, along with the susceptibility rates and mortality rates was carried out for the South Asian countries. The only two countries able to hold the disease at a detectable level are Nepal and Bhutan. The transport and communication ways available, and the geographical barriers were taken into account to see its effects on the spread of the disease. There has been a marked gender disparity in COVID-19 cases across the area (Velavan *et.al*., 2020). The reasons could be multiple like higher level of exposure, severity of disease, access to testing etc., but men constitute the majority of the cases. We also took into account the different blood groups in the countries and checked the susceptibility rate towards the infection and if there was a direct correlation between the blood type antibodies and the mortality rate. The death rates were accelerated in co morbid cases, where pre-existing medical conditions like hypertension, diabetes, and heart diseases were present. The role of specific antihypertensive medications like angiotensin-converting enzyme inhibitors (ACEIs) and angiotensin receptor blockers (ARBs) was studied because it was linked to angiotensin-converting enzyme 2 (ACE2), a co receptor for viral entry for SARSCoV-2 with increasing evidence that it has a protracted role in the pathogenesis of COVID-19 (Rothan *et.al*., 2020). The ACE2 was broadly expressed in in the gastrointestinal system, heart, and kidney with more recent data identifying expression of ACE2 in type II alveolar cells in the lungs. The food habits across the South Asian countries are very similar, the staple food being rice, flatbreads, pulses and lentils, starchy vegetables, and sea food. The use of spices and herbs like turmeric, chilies, curry leaf, coriander, galangal, cinnamon, cumin, cardamom, fenugreek, pepper, and cloves are widespread and popular. Climatic factors might have a good role to play in the spread of the COVID-19 disease across the areas. Sensitivity analysis results show that the population density, intra-provincial movement play a direct role at the rate of infectivity (Singh *et.al*., 2020). Thus, showing that areas with low values of wind speed, humidity, and solar radiation exposure has a high rate of infection because it well supports the survivability of the virus. It was observed that even areas with low population density but with relative humidity and/or low wind, speed the rate of infectivity is higher. A comparative study was also done to see the preparedness of the countries to tackle the pandemic in terms of the health support available with respect to the affected patients, the medications used for treatment, and lastly the economic conditions of the country, their respective GDP/capita, inflation rates and the proportion of poverty prevalent across the countries.

Our main objective was to take into account the various quantitative and qualitative factors like the geographical and climatic diversity, the economic and healthcare system of the South Asian countries, the inherent physiological conditions of the people and the impact of these factors on the spread of Covid-19, and thereby calculate the infectious rate and fatality of the disease across the South Asian countries.

## 3. Materials and Methods

### 3.1. Collection of data

#### 3.1.1. Population of the South Asian Countries

Several scientific models have proved that the spread of any communicable disease is directly dependent on the population graph of any country (Kopec et.al., 2010). Novel corona virus being a global concern right now, the main source of spread of the infection is the transmission via airborne droplets (Morawska *et.al*., 2020). The transmission of expiratory droplets highly depends on the distance between two people and thus population is an important parameter which can play a key role in the spread of Covid-19 (Liu *et al*., 2017). Thus, we have collected the respective total populations of the seven south Asian countries from www.worldometer.info.

#### 3.1.2. Population Density across South Asian Countries

Scientists suggest that the high population densities are the major driving factor behind the spread of Covid-19 (Rocklöv et.al.,2020). Hence, the population densities of the respective south Asian countries were collected from www.worldometer.info.

#### 3.1.3. Population tested for Covid-19

According to World Health Organization, testing is the principal factor for controlling the spread of SARS-CoV-2 (World Health Organization, March 2020). The incubation period for the novel corona virus ranges from 4-14 days. Hence if the test frequency is increased, then both symptomatic patients as well as asymptomatic carriers can be easily detected, thereby controlling spread of the disease to a certain extent (Beeching et.al., 2020). The data for population tested for the different countries was collected from respective reliable online sources. Percentage is calculated using the formula

(Total number of people tested/ Total population of a country) x 100

#### 3.1.4. Covid-19 positive cases across the South Asian Countries

After Covid-19 was declared a pandemic by WHO with 2,00,000 affected patients and 8000 deaths over 160 countries, the virus was observed to spread very rapidly such that within 2 weeks from the first cases diagnosed (1000 patients), the number of positive cases exceeded 4,600 and reached 30,000 on 18^th^ of March, 2020 (Spinelli et.al., 2020). Scientific research and mathematical model claim that isolation and contact tracing can be used to control Covid-19 outbreaks (Ph.D. Joel Hellewell, et al). Thus, higher rate of diagnosis of positive covid-19 cases and their isolation can be a major deciding parameter. The formula used for calculating percentage of positive Covid-19 cases diagnosed is

(Total number of positive cases/ Total number of people tested) x 100

#### 3.1.5. Covid-19 recovered cases across South Asian Countries

Having a clear understanding about the recovery time and rate of a disease is of utmost importance since that information can help in tackling the disease. If both the occurrence and the recovery time of a disease like Covid-19, is high, then the prevalence of the disease in densely populated countries like India, Bangladesh, Pakistan, is likely to put an extra burden on the healthcare facilities as well as the economic and social systems. Such insight can help the government to properly plan strategies like requirement of hospitals, doctors, medical stuff and equipments and make different social and economic policies that will counter the disease and its adverse socio-economic effects (Barman et.al., 2020). A major variation in the number of testing carried out was observed in the South Asian Countries with respect to other countries like USA, Germany, South Korea, etc. (Dhillon et.al., 2020). From our study, it was evident that as the number of testing increased, the number of recovered individuals also increased simultaneously.

#### 3.1.6. Covid-19 death cases across South Asian Countries

The total number of Covid-19 cases of South Asia on April 1, 2020 was 3,649 which increased to 62,895 on May 1, 2020 and crossed 2,00,000 on 21^st^ May, 2020. Correspondingly, the total death cases increased from 71 on April 1, 2020 to 1,772 on May 1, 2020 and around 5,000 on 21^st^ May, 2020. Amongst the South Asian countries, as of 21^st^ May, 2020, no mortality was reported in Bhutan and only 3 death cases were announced in Nepal (Khafaie et.al., 2020). India, on the other hand, reported the maximum number of confirmed Covid-19 cases (>1,00,5000) and highest number of deaths (>3,500) as of 21^st^ May, 2020. South Asia, being one of the most densely populated and poorest regions in the world with substandard healthcare facilities, it reported an increasing number of casualties. However, due to strict lockdown, high youth population and low testing frequency, the mortality was not as high as expected.

#### 3.1.7. Percentage of population tested for Covid-19 across South Asian countries

No country is aware of the total number of people infected with Covid-19, however, all we know is the infection status of the ones who have been tested. This means that the number of confirmed cases is directly proportional to the number of tests carried out in the country. Testing reveals the percentage of population affected and indicates how seriously a country is affected by the pandemic. Without testing there is no data and we would not understand which countries are doing well in countering the, disease and which are just under reporting cases and deaths. When there is limited testing, many cases are likely to be missed and a high positive rate of infection hints that the number of confirmed cases is only a small fraction of the true number of individuals affected (Bhadra et.al., 2020). Thus, percentage tested is given by –

Percentage of population tested for Covid-19

= (Total no. of persons tested/ Total population) x 100

This value is an important tool to fight to slow and reduce the spread and impact of this deadly virus. A very good example was cited by the state of Kerala in India, where the government prioritized systematic testing and widespread contact tracing. Thereby, they implemented uncompromising control and social distance measures, which curbed the spread to a massive extent.^[14^

#### 3.1.8. Percentage of Covid-19 Positive Cases across South Asian Countries

The initial cases reported in the South Asian countries were primarily the travelers who returned from other Covid affected countries like China, USA, etc. Gradually the positive cases increased in South Asia and by April 21, 2020, 31565 individuals were affected of which 5,526 recovered and 901 succumbed. As of April 21, India had the highest percentage of positive cases (59.1%), followed by Pakistan (29.2%), Bangladesh (10.7%) and Sri Lanka (1%). The spread of the disease was very rapid in India and Pakistan and not as much in Bangladesh, Sri Lanka, Nepal and Bhutan (Bhadra et.al., 2020). A probable reason for such disparity could be a high proportion of the economically disadvantaged class living in crowded slums, where social distancing and basic hygiene are farfetched concepts. The percentage tested was given by-

Percentage of Covid-19 positive cases

= (No. of positive cases/Total number of persons tested) x 100

#### 3.1.9. Percentage of Covid-19 Recovered Cases across South Asian Countries

Amongst the South Asian Countries, a maximum recovery rate of 48.18% was observed in India as of 31^st^ May, 2020. The recovery rate is given by –

Percentage of Covid-19 recovered cases

= (Recovered/No. of Positive Cases) x 100

In India, it was found that, an average of 25 days under treatment was required to fully recover from Covid-19. These recovery rates and time are important because the government will accordingly make strategies for public health such as arrangements of doctors, nurses, medical stuff, isolation beds, etc. ^(^Barman et.al., 2020).

#### 3.2.0. Covid-19 Case fatality rate (CFR)/Mortality rate across South Asian Countries

The mortality or case fatality rate (CFR) is the measure of the virulence of a pathogen that is the ability of it to cause fatal damage to the host. For an infectious disease, CFR is defined as the percentage of positive cases that result in death. The proportion of death cases indicates the severity of the disease and thus knowing the CFR is important for taking appropriate preventive measures and setting priorities for public health (Khafaie et.al., 2020). As of April 21, 2020, the percentage of reported death or CFR was highest in Bangladesh (55.84%) followed by India (15.32%), Pakistan (8.5%) and Sri Lanka (6.54%). The Case Fatality Rate was calculated as follows –

Case Fatality Rate or Mortality percentage

= (No. of deaths/No. of positive Cases) x 100

#### 3.2.1. GDP Per Capita (in US Dollars) across SA Countries

The economic risk of Covid-19 was geo-spatially evaluated to determine the vulnerability and resilience of the local economy to the shock of the pandemic. It was found that the South Asian Countries were particularly in the high-risk zone when compared to the US and European countries. Assessing the economic condition is important for bringing about changes in the socio-economic policies that are to be implemented in the public health domain (Santosh, K.C., 2020). To understand the economic condition of the South Asian Countries, we took into consideration the GDP per capita (in US Dollars) of the countries of the year 2018.

#### 3.2.2. Percentage of Population below poverty line in South Asian Countries

“Covid-19 does not discriminate.” – is a dangerously illusive myth because it sidelines the increased vulnerability of the socially and economically deprived class. For people belonging to low socio-economic status (SES), there are a number of factors that increase their exposure to Covid-19. First, these people live in over-crowded houses with poor hygienic conditions, where the norms of social distancing cannot be applied. Second, these people are employed in occupations that do not provide opportunities to work from home. So, they might have to avail various forms of crowded public transport to reach their workplace, which has often resulted in them contracting the infection. Third, people belonging to low SES groups have unreliable work place and erratic income, which got aggravated in the Covid situation. Such financial uncertainty harms mental health, escalating stress levels that in turn weakens the immune system, increasing the vulnerability to Covid. Finally, people belonging to low SES are likely to approach the healthcare services at an advanced stage of the disease resulting in poorer health outcomes (Patel J. A. et. al., 2020). To compare the poverty levels of the South Asian countries, we took into account the percentage of population below poverty line in these countries.

#### 3.2.3. Help Support available with respect to affected patients (Beds/1000)

The Covid-19 mortality was found to be inversely proportional to the effectiveness of the government to formulate and implement sound policies in the healthcare sector to tackle the emergency. This includes provision of adequate hospital beds, medical facilities, critical care, ventilators for the high-risk patients, etc. (Santosh, K.C., 2020). The number of beds per thousand available in each of the South Asian countries were noted and a comparative study was carried out.

#### 3.2.4. Average temperature and Relative humidity across South Asian Countries

A relevant question that arise is if the transmission of Covid-19 is dependent on climatic factors like temperature and humidity that is, is it less transmissible in hot and humid climates. Studies have shown that regions with lower mean temperature and specific humidity reported significant community outbreaks. It was reported that SARS COV I had a lower survival rate at higher temperature and humidity (Rashed et.al., 2020). To find out how the climatic conditions of the South Asian Countries determined the viral transmission of SARS COV II, we acquired the Average temperature and Relative humidity data and carried out a comparative study (Gupta et.al., 2020).

#### 3.2.6. Data analysis using R Studio

After collecting the epidemiological, economic and climatic data we carried out a comparative study by finding and analyzing the Karl Pearson’s Correlation Coefficient for the various factors.

## 4. Results

### 4.1. Population of the South Asian Countries

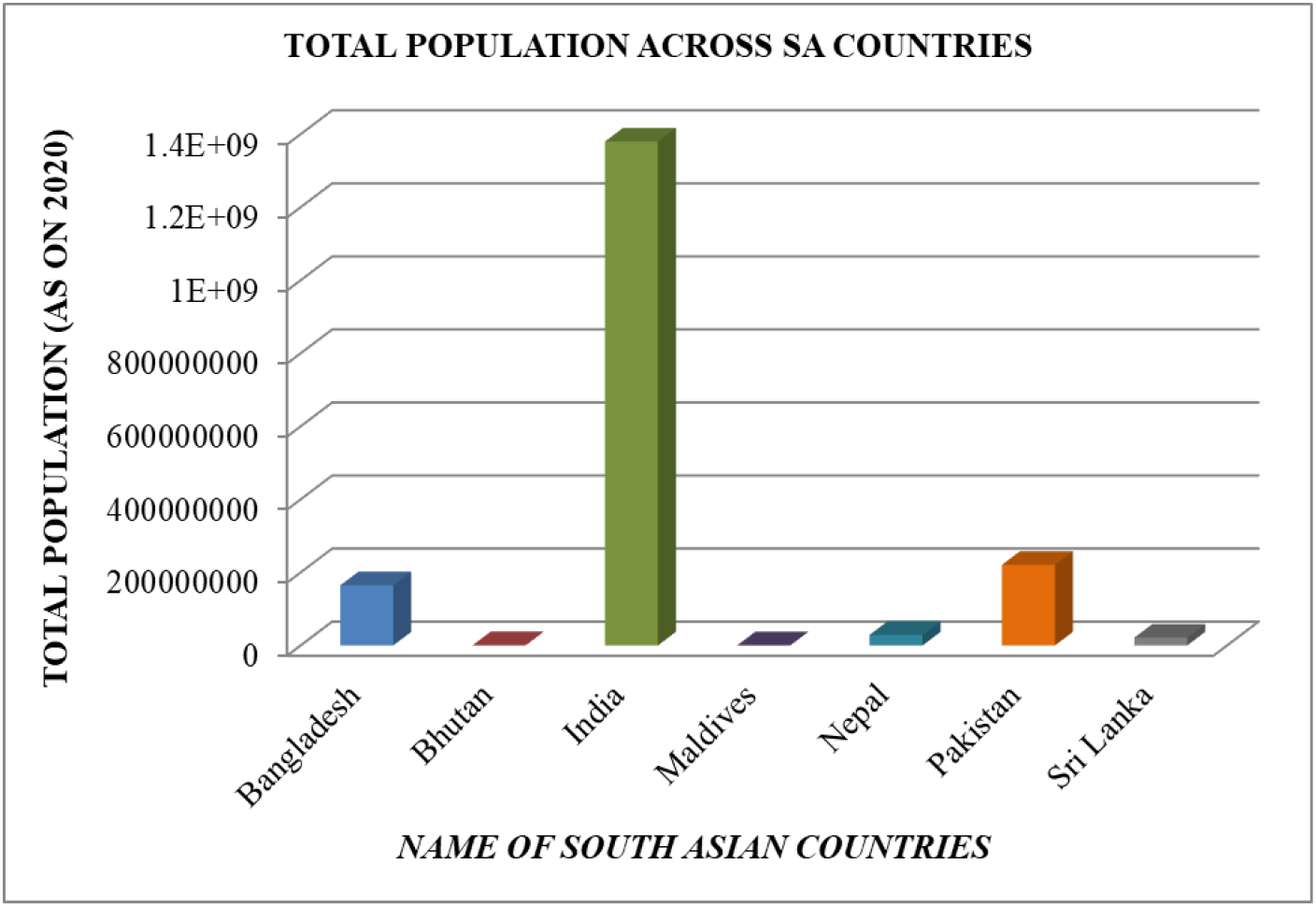

The above graph shows that range of total population of the seven South Asian countries under our study, the blue color representing Bangladesh, red representing Bhutan, green representing India, purple representing Maldives, turquoise representing Nepal, orange representing Pakistan and grey representing Sri Lanka. From the graph, it can be concluded that India has the highest range of total population followed by Pakistan, Bangladesh, Nepal, Sri Lanka, Bhutan and Maldives having the least population of all.

#### Inference

Therefore, from here we can infer that, total population of a country can play a major role in the spread of the infection of Covid-19.

### 4.2. Population Density across South Asian Countries

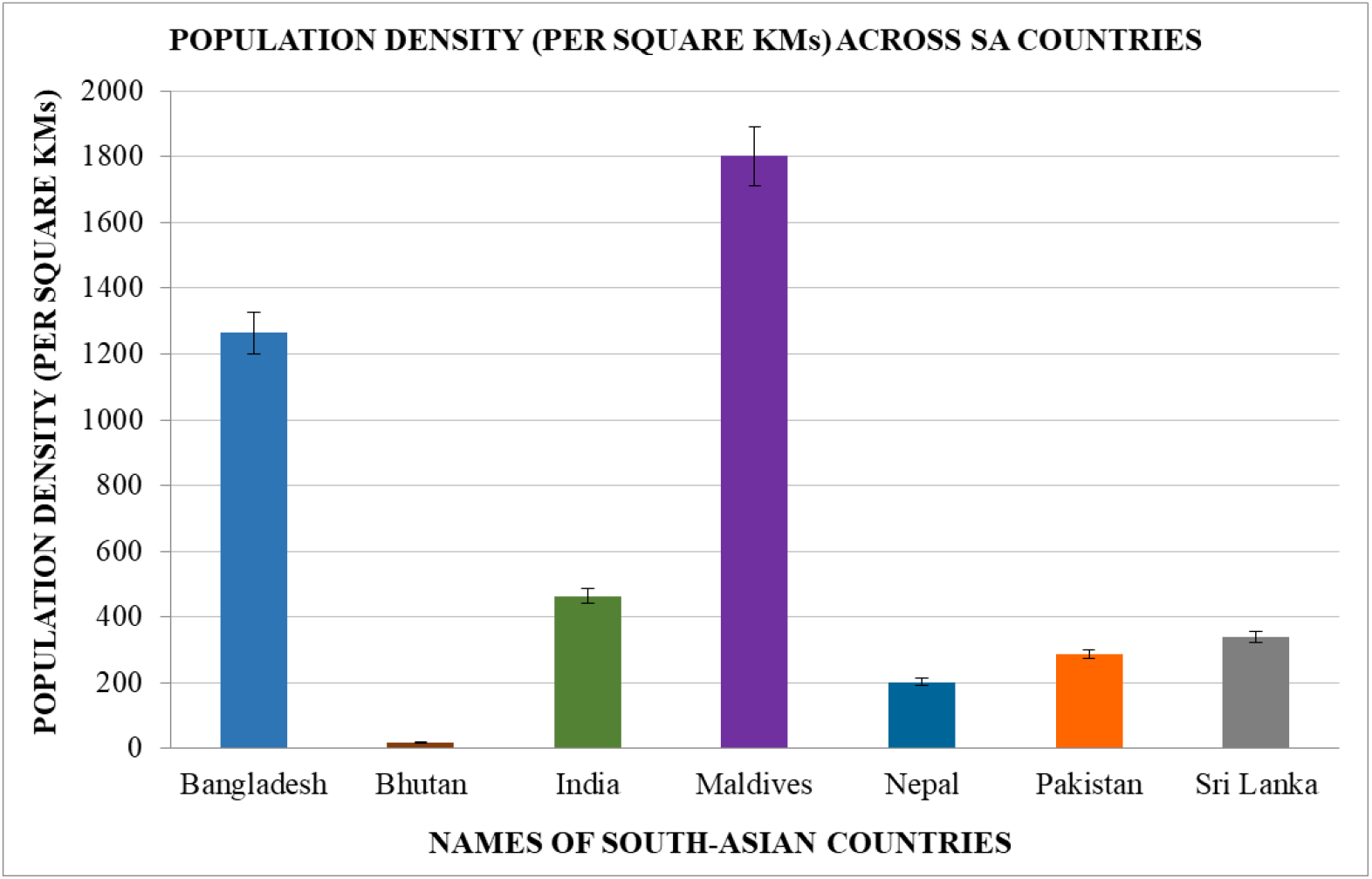

The above graph shows the range of population density of the seven South Asian countries under our study, where the highest population density is that of Maldives, followed by Bangladesh, India, Sri Lanka, Pakistan, Nepal and Bhutan.

#### Inference

Therefore, from the above data we can infer that population density of a country might play an important role in the spread of the infection of Covid-19, since, a high population density is indicative of the fact that the population of that country is relatively high in comparison to the size of the country.

### 4.3. Population tested for Covid-19

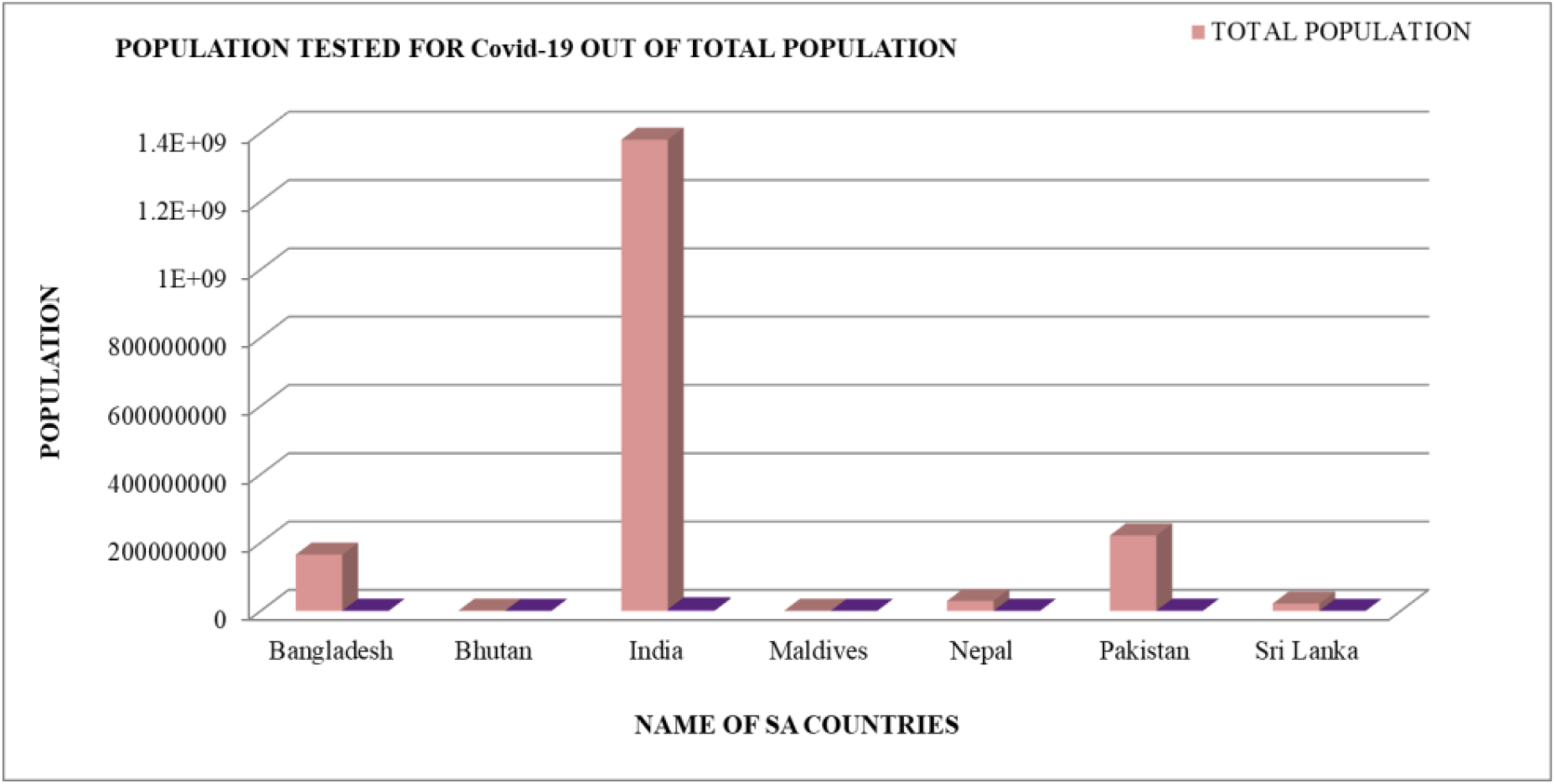

The above graph shows the total number of people tested for Covid-19 out of the total population of the seven South Countries respectively, of our study. The graph gives a clear comparison between the total number of people tested with respect to the total population, where, the peach color represents the total population of the seven countries and purple represents the total number of people tested for Covid-19 in each of the seven countries.

#### Inference

So, from the above graph we can infer that a high number of people tested out of the total population gives a clear idea about the spread of the infection of Covid-19 in each of the seven countries.

### 4.4. Covid-19 positive cases across the South Asian Countries

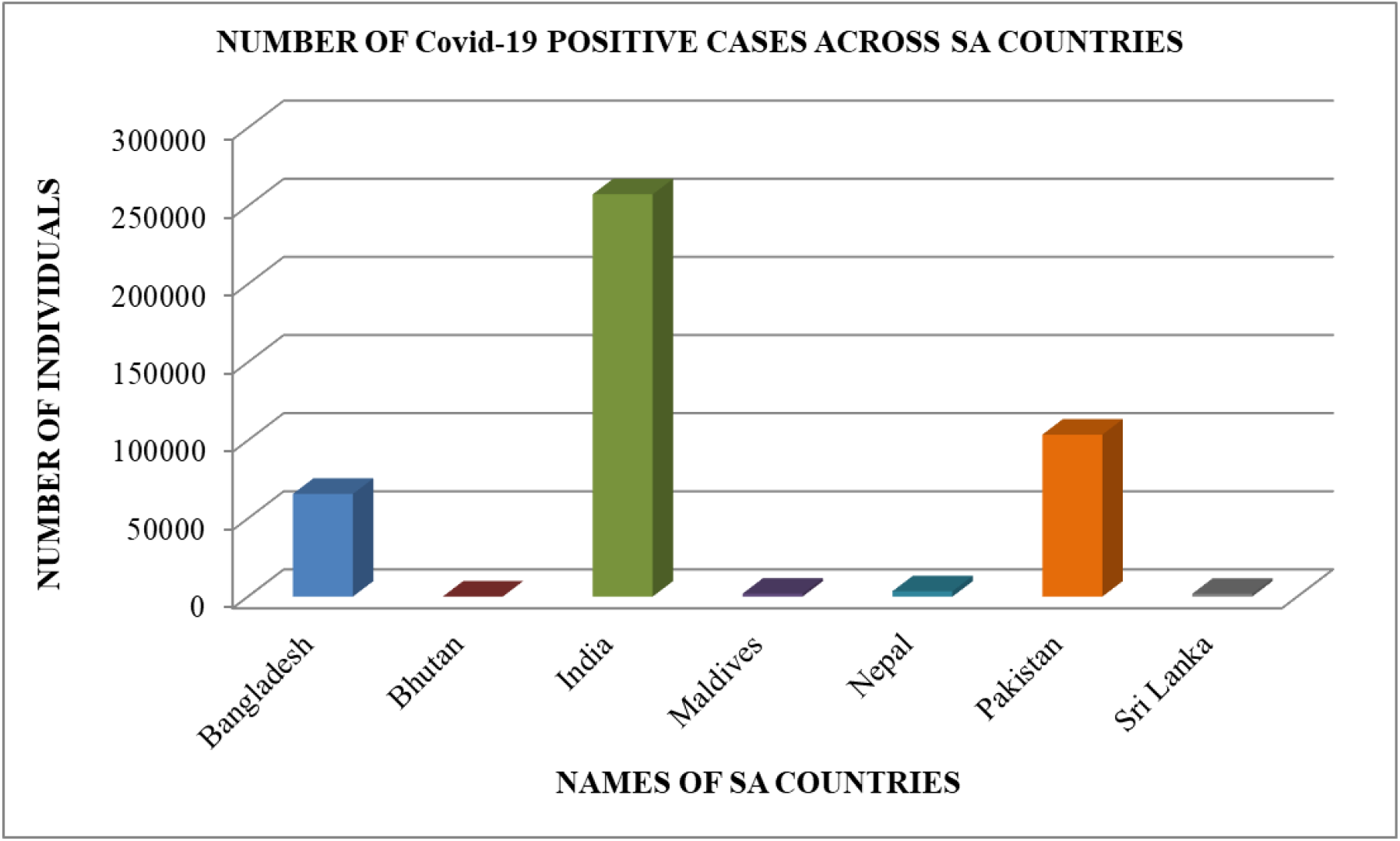

The above graph shows the number of Covid-19 positive cases in each of the seven South Asian countries, where, the highest number of positive cases is observed in India followed by Pakistan, Bangladesh, Nepal, Bhutan Sri Lanka and Maldives.

#### Inference

Hence, from the above graph, we can infer that the highest spread of the infection is observed in India as it recorded the highest range of positive cases, followed by Pakistan and Bangladesh.

### 4.5. Covid-19 recovered cases across South Asian Countries

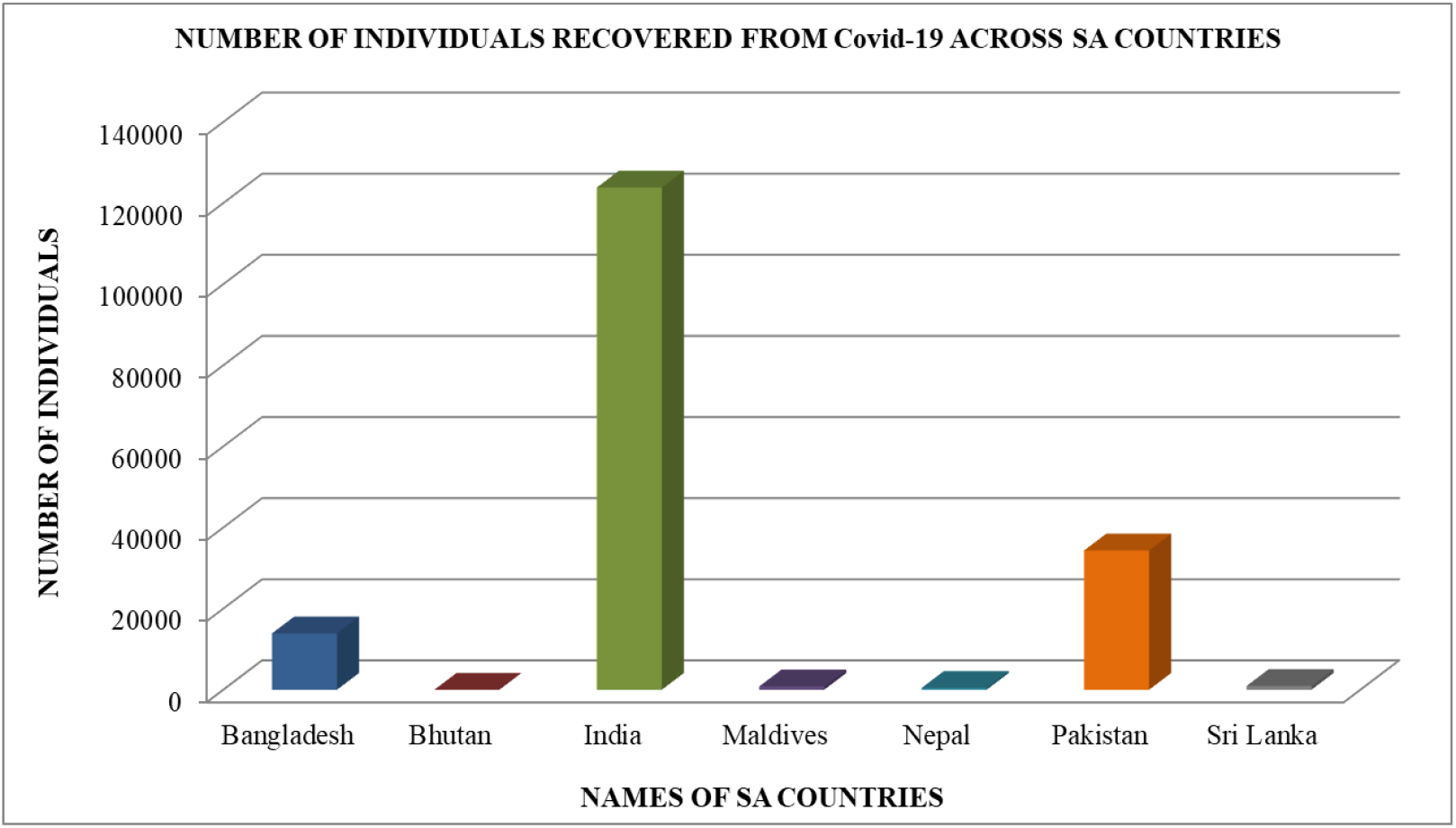

The above graph shows the range of total number of Covid -19 recoveries in each of the seven South Asian Countries, where the highest range was obtained by India, followed by Pakistan and Bangladesh.

#### Inference

Hence, from the above graph, we can infer that the highest number of recoveries in the time of our study was observed in India. The next country to show significant number of recoveries was Pakistan followed by Bangladesh.

### 4.6. Covid-19 death cases across South Asian Countries

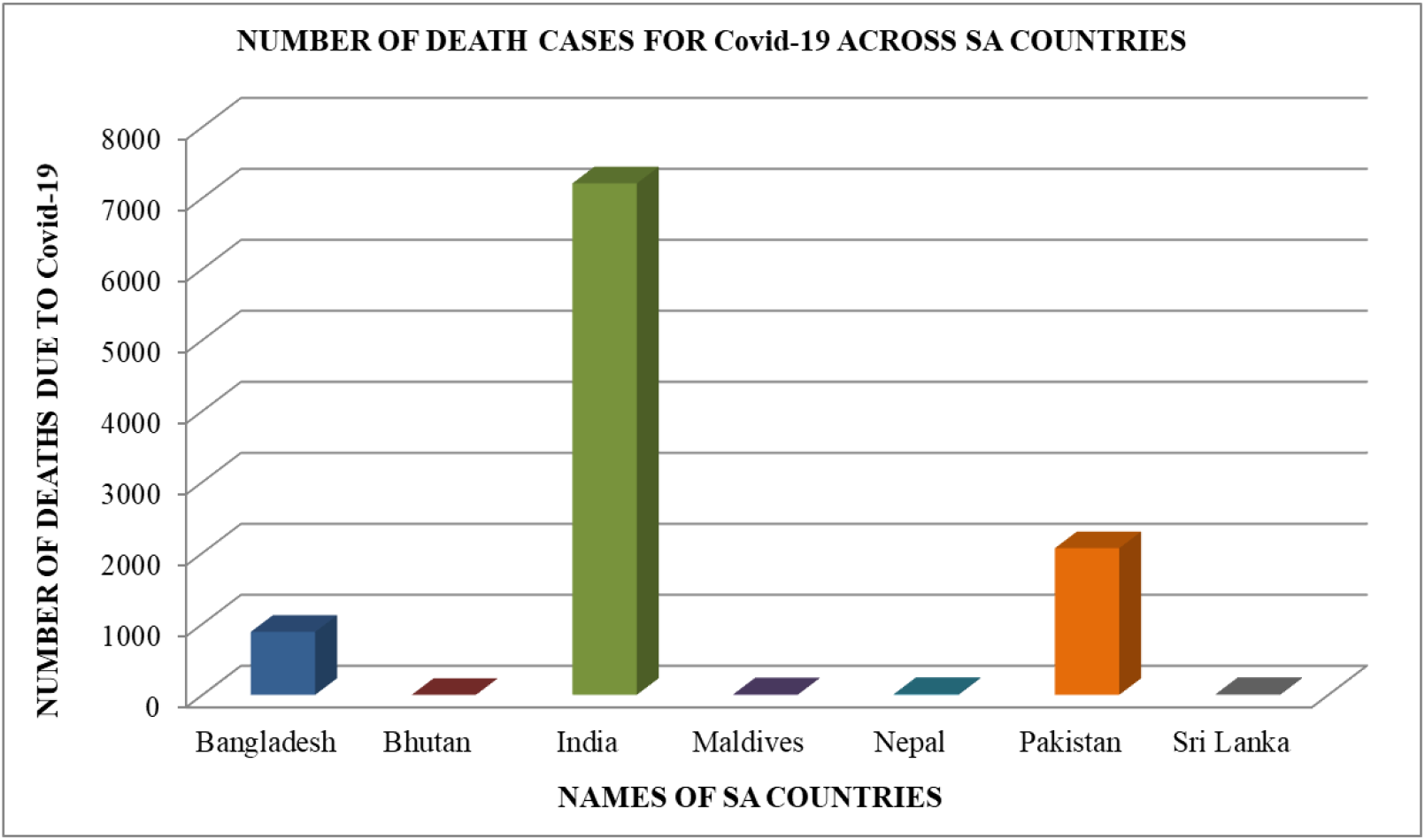

The above graph shows the range of total number of Covid -19 death cases within the time of our study in each of the seven South Asian Countries. The maximum range was obtained for India, followed by Pakistan and Bangladesh.

#### Inference

Hence, from the above graph, we can infer that the highest number of death cases in the time of our study was observed in India. The next country to show significant number of death cases was Pakistan followed by Bangladesh.

### 4.7. Percentage of population tested for Covid-19 across South Asian countries

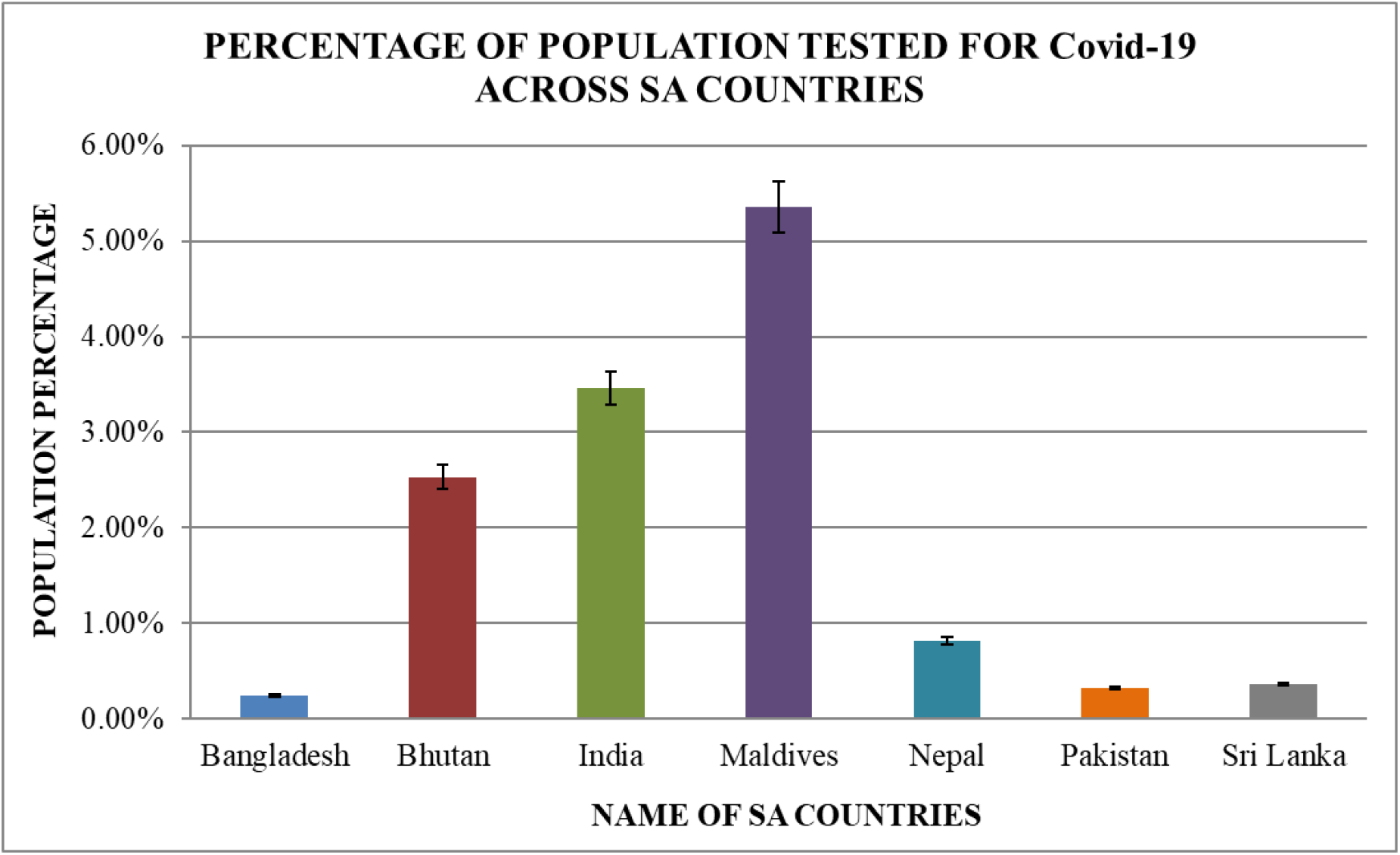

The percentage of population tested was calculated using the formula – Percentage of population tested for Covid-19

= (Total no. of persons tested/ Total population) x 100

The above graph gives an account of the percentage of population tested for Covid-19, in each of the seven South Asian Countries within our time of study. The maximum range was observed for Maldives followed by India, Bhutan, Nepal, Sri Lanka, Pakistan and lastly Bangladesh.

#### Inference

Hence, from the above graph, we can infer that the highest range of population tested for Covid-19, was observed in Maldives closely followed by India, Bhutan and Nepal, Sri Lanka while in Pakistan and Bangladesh very low percentage was observed which indicated towards a possibility of under testing.

### 4.8. Percentage of Covid-19 Positive Cases across South Asian Countries

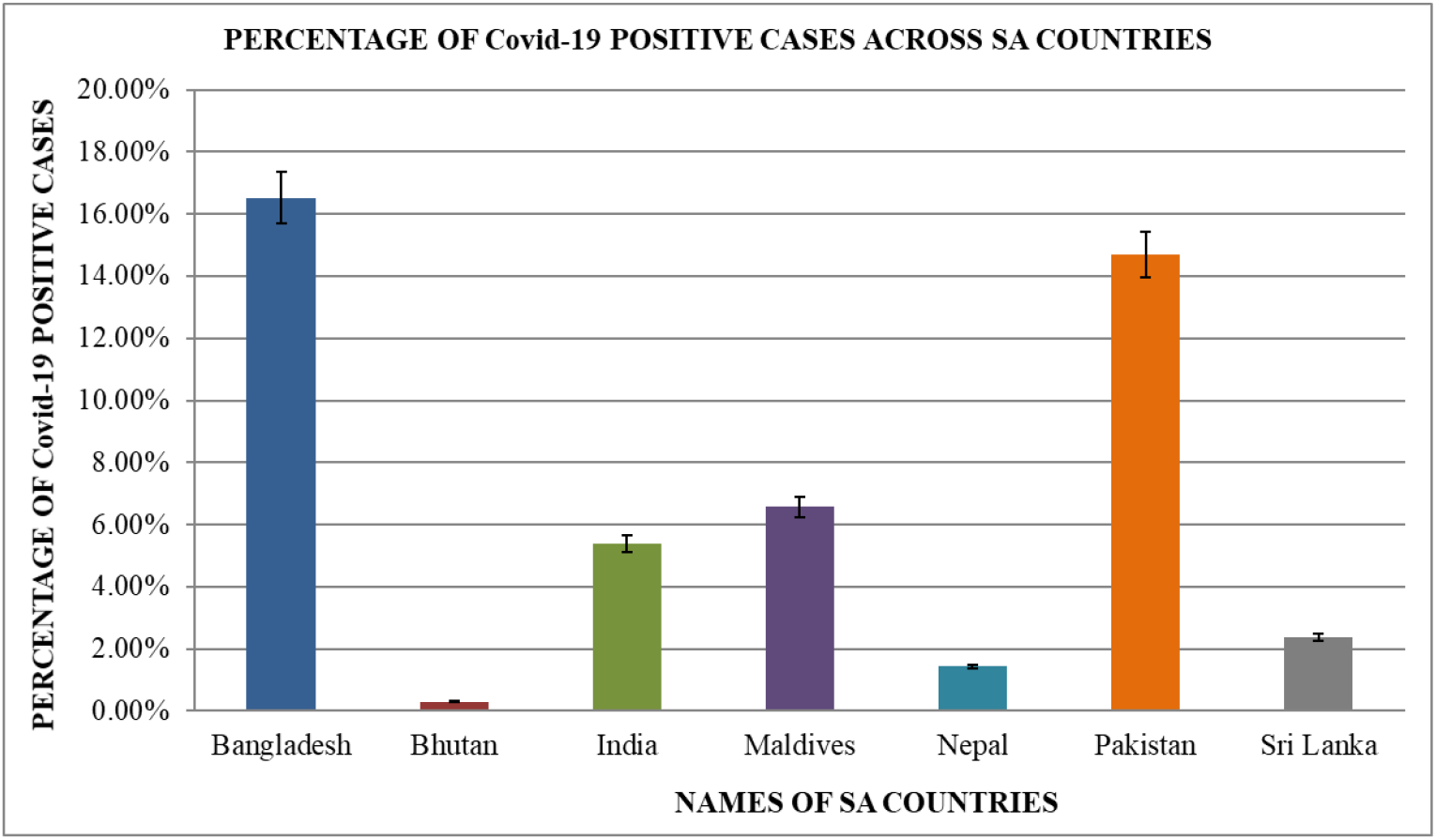

The percentage of Covid-19 positive cases was calculated using the formula – Percentage of Covid-19 positive cases

= (No. of positive cases/Total number of persons tested) x 100

The above graph gives an account of the percentage of Covid-19 positive cases in each of the seven South Asian countries within our time of study. The maximum range was observed for Bangladesh followed by Pakistan, Maldives, India, Sri Lanka, Nepal and Bhutan.

#### Inference

Hence, from the above graph we can infer that maximum number of individuals out of the total number tested, tested positive for Covid-19 in Bangladesh and Pakistan, while the range was relatively lower for Maldives and India and significantly less in Nepal, Bhutan and Sri Lanka.

### 4.9. Percentage of Covid-19 Recovered Cases across South Asian Countries

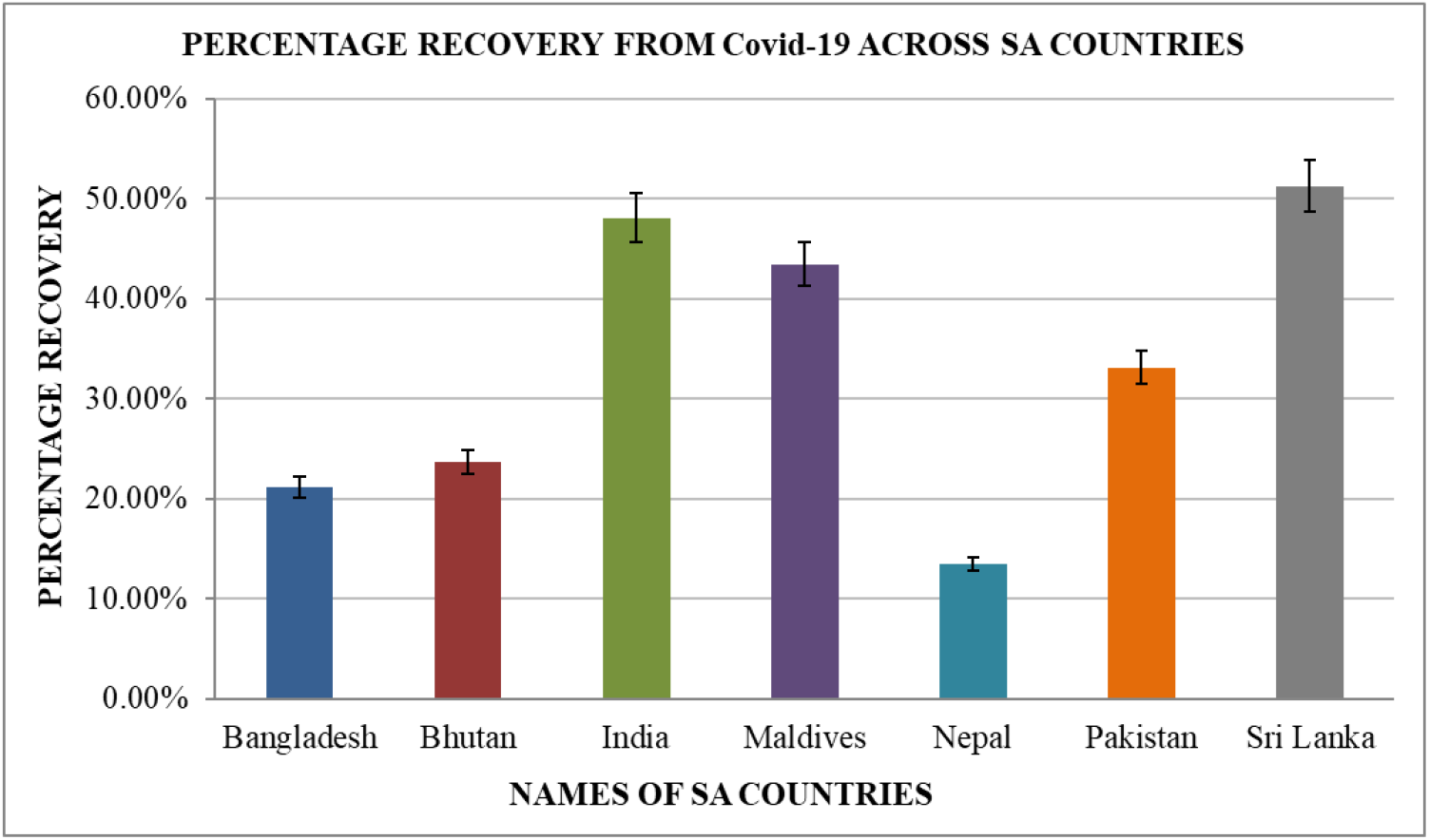

The recovery rate is given by – Percentage of Covid-19 recovered cases

= (Recovered/No. of Positive Cases) x 100

The above graph gives an account of the percentage of Covid-19 recovered cases in each of the seven South Asian countries within our time of study. The maximum range was observed for Sri Lanka, followed by India, Maldives, Pakistan, Bhutan, Bangladesh and Nepal.

#### Inference

Hence, from the above graph we can infer that the maximum recovery rate was observed in case of Sri Lanka, closely followed by India, Maldives and Pakistan while it was comparatively less for Bhutan, Bangladesh and Nepal.

### 4.10. Covid-19 Case fatality rate (CFR)/Mortality rate across South Asian Countries

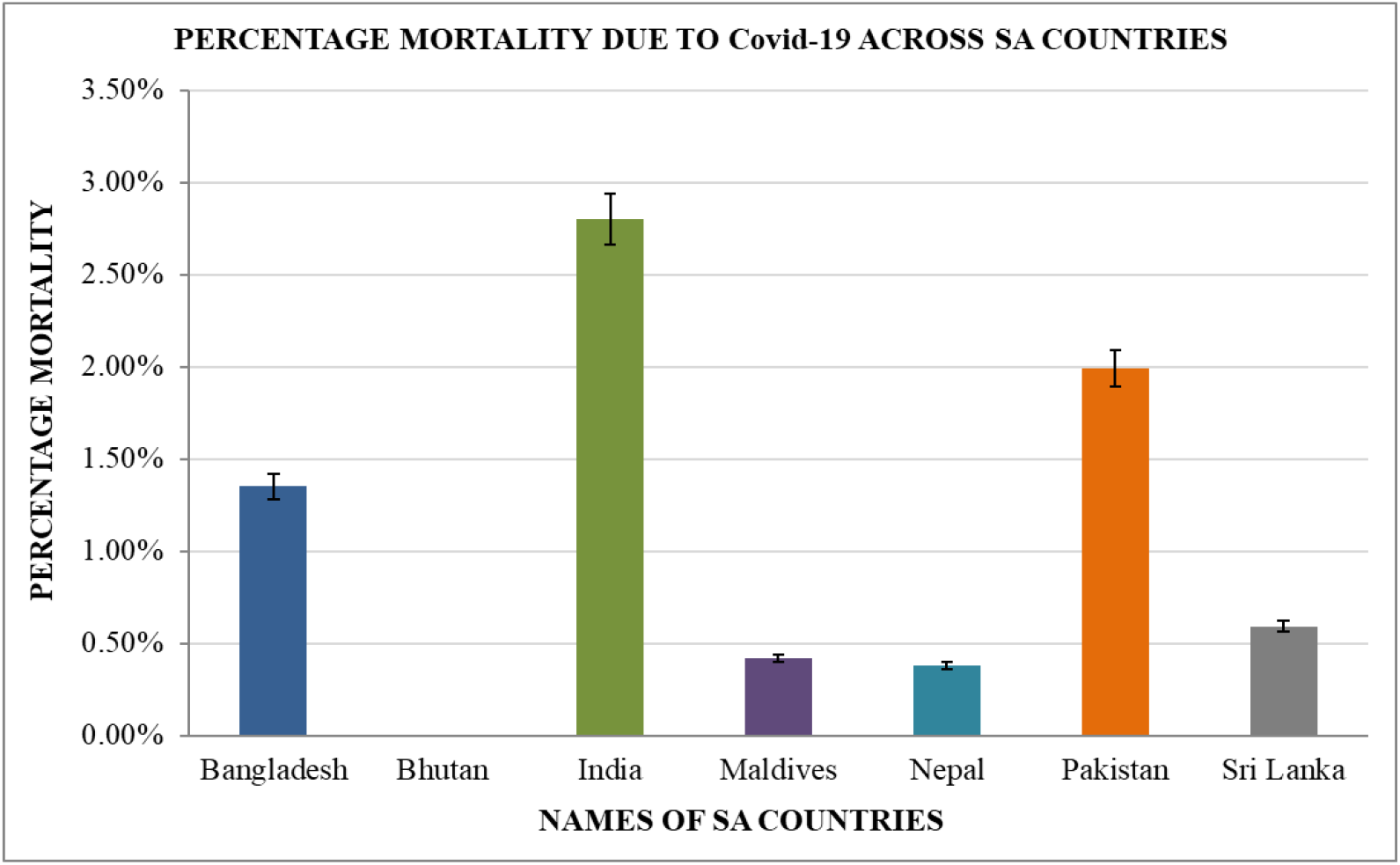

The Case Fatality Rate was calculated as follows –

Case Fatality Rate or Mortality percentage

= (No. of deaths/No. of positive Cases) x 100

The above graph gives an account of the Case Fatality Rate due to Covid-19, in each of the seven South Asian countries within our time of study. The maximum range was observed for India, closely followed by Pakistan, Bangladesh, Sri Lanka, Maldives and Nepal. Bhutan showed no observation in this regard.

#### Inference

Hence, from the above graph we can infer that the maximum fatality rate was observed in case of India, followed by Pakistan, Bangladesh, Sri Lanka, Maldives and Nepal. In Bhutan however, no death due to Covid was observed within our time of study, as a result which, the case fatality rate could not be calculated.

### 4.11. GDP Per Capita (in US Dollars) across SA Countries

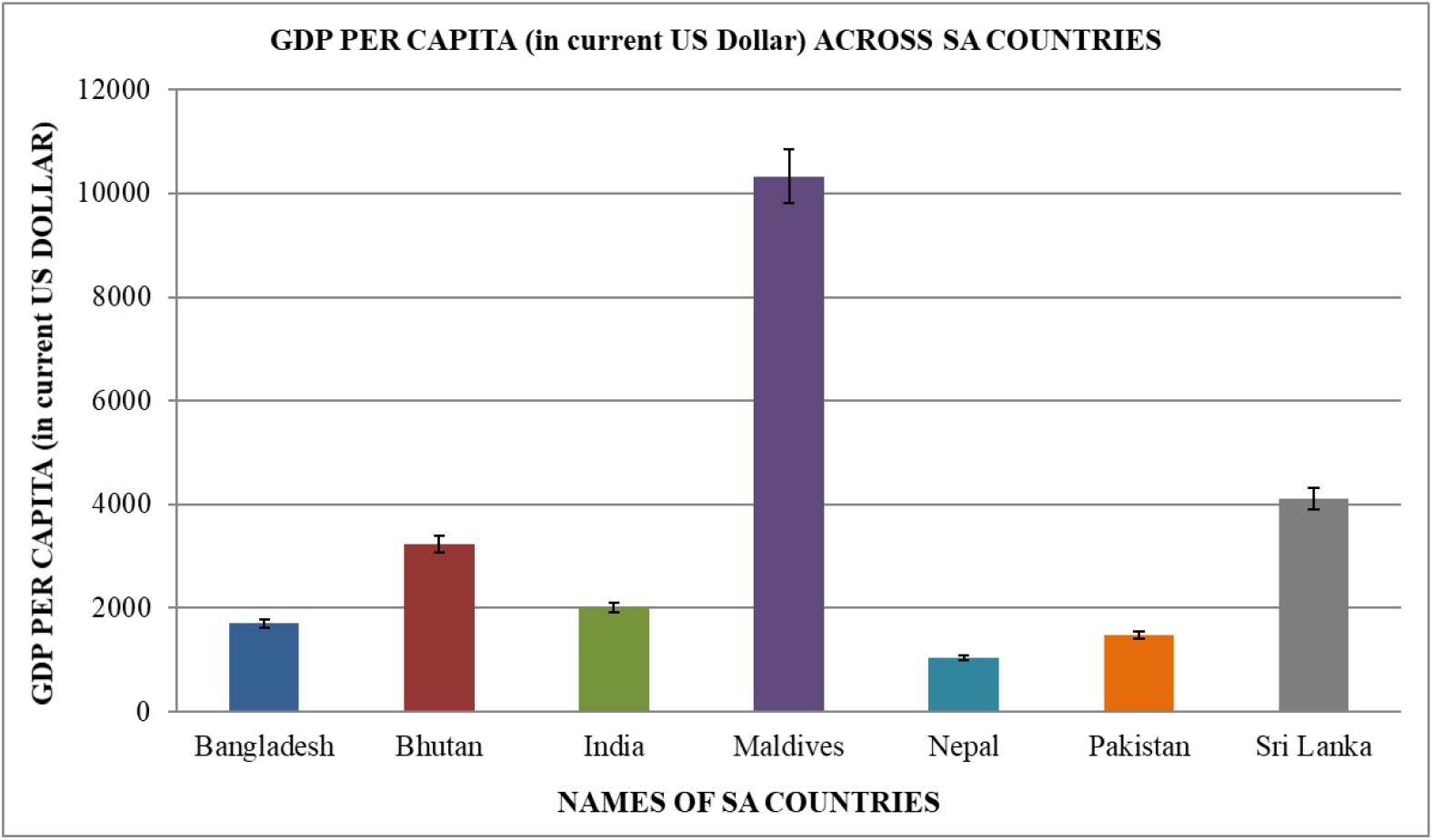

The above graph gives an account of the GDP Per Capita (in US Dollars) in each of the seven South Asian countries. The maximum range was observed for Maldives, followed by Sri Lanka, Bhutan, India, Bangladesh, Pakistan and Nepal.

#### Inference

Hence, from the above graph we can infer that the maximum range for GDP Per Capita (in US Dollars) was observed for Maldives followed by Sri Lanka and Bhutan, thereby indicating towards the increased probability of efficient dealing of the Covid-19 pandemic by these countries.

### 4.12. Percentage of Population below poverty line in South Asian Countries

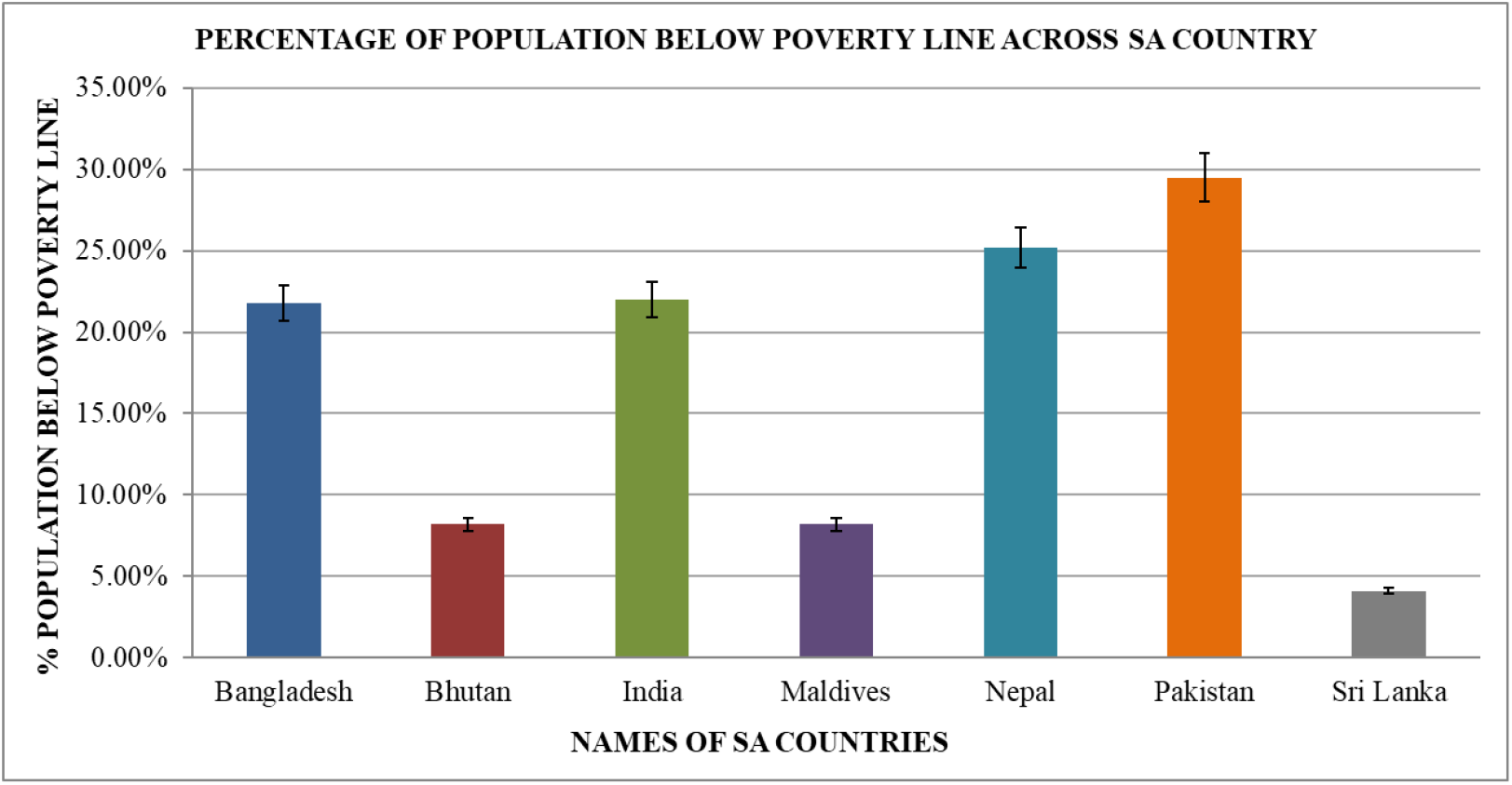

The above graph gives an account of the Percentage of Population below poverty line in each of the seven South Asian countries. The maximum range was observed for Pakistan, followed by Nepal, India and Bangladesh, Bhutan and Maldives, and lastly Sri Lanka.

#### Inference

Hence, from the above graph we can infer that the maximum percentage of population below the poverty line was observed for Pakistan, indicating towards an increased level of poverty in the country. Pakistan was closely followed by Nepal, India and Bangladesh, while the observations for Bhutan, Maldives and Sri Lanka were comparatively in the lower range.

### 4.13. Help Support available with respect to affected patients (Beds/1000)

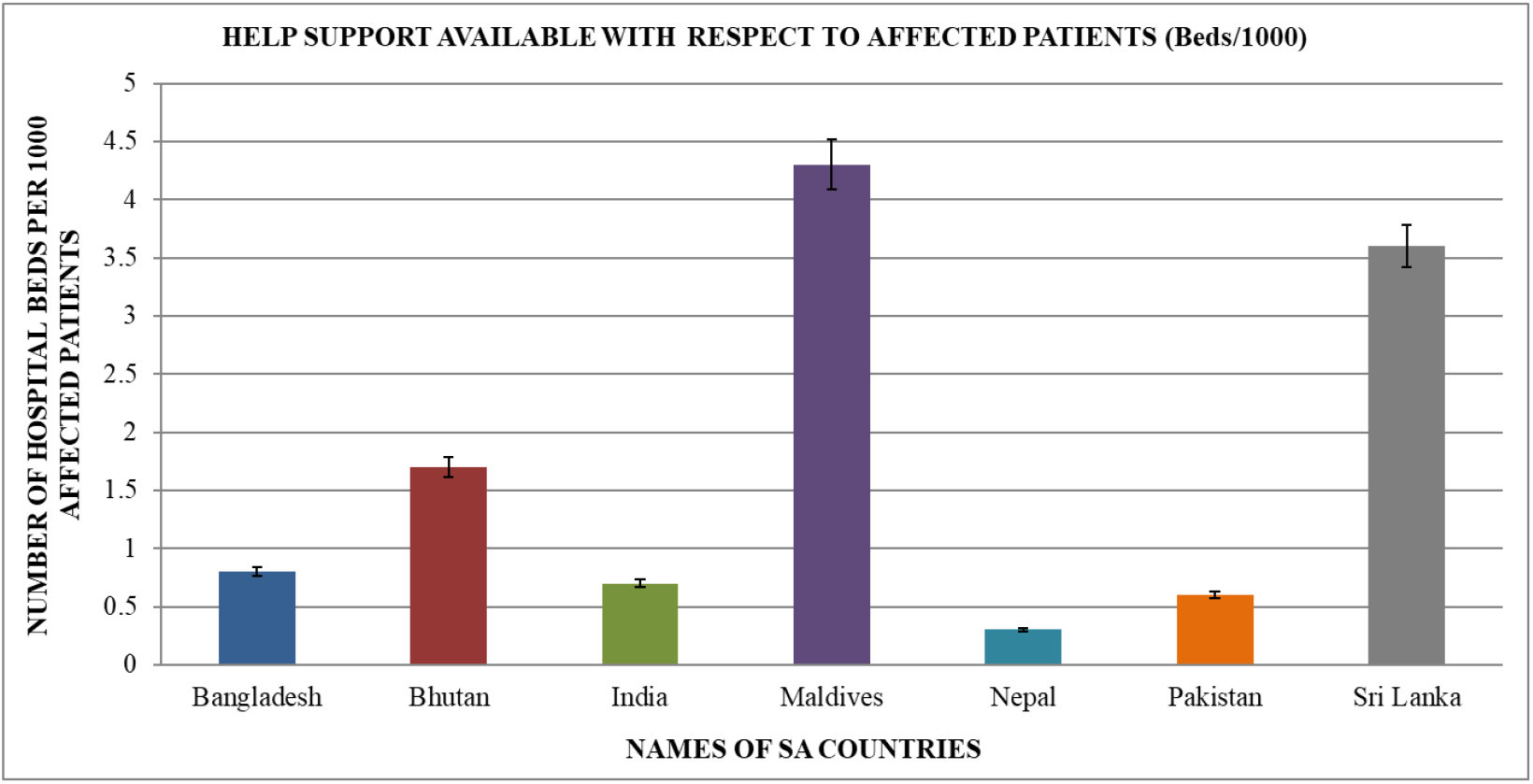

The above graph gives an account of the Help Support available with respect to affected patients (Beds/1000) in each of the seven South Asian countries. The maximum range was observed for Maldives closely followed by Sri Lanka. The data of the countries were significantly lower and moderate ranges were observed for Bhutan, Bangladesh, India, Pakistan and Nepal.

#### Inference

Hence, from the above graph we can infer that Maldives has the best help support available with respect to affected patients (Beds/1000) followed closely by Sri Lanka out of the seven SA countries. The health infrastructure of these countries therefore show great possibility for dealing efficiently with the Covid-19 pandemic.

### 4.14. Average temperature and Relative humidity across South Asian Countries

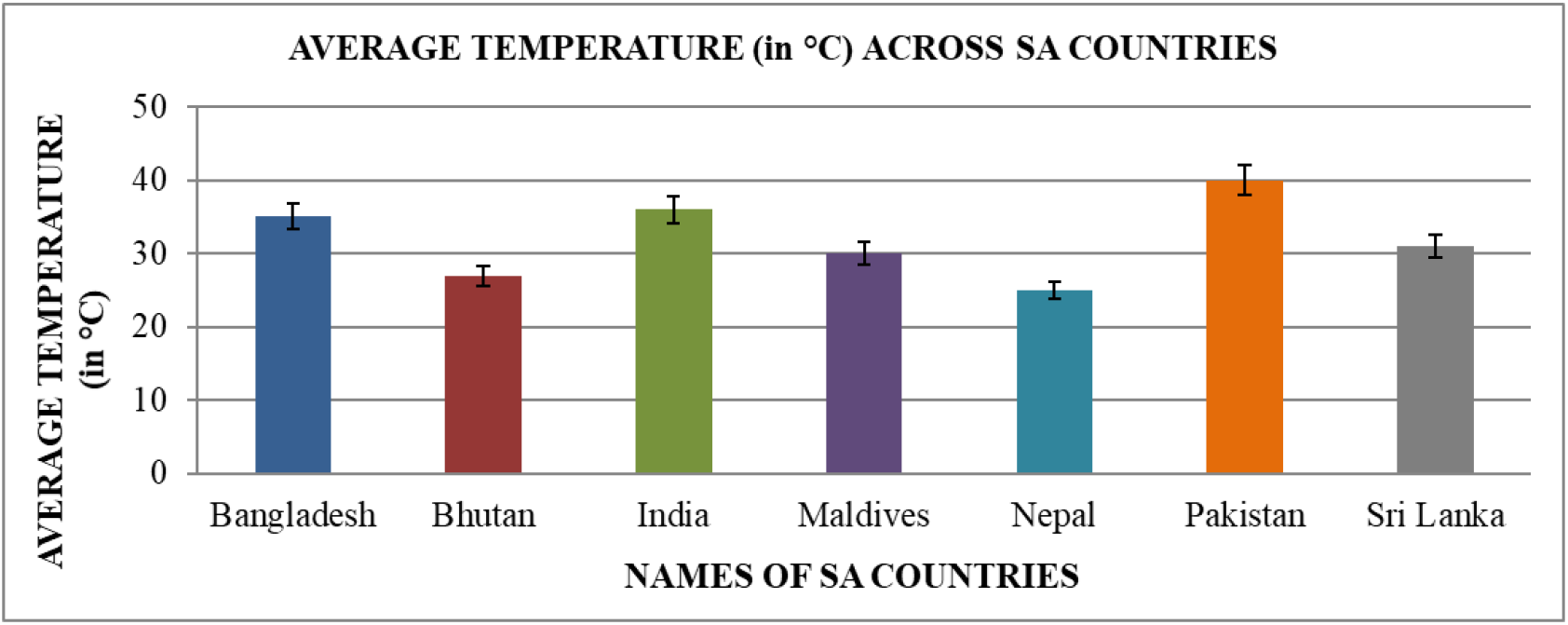

The above graph gives an account of the Average temperature in each of the seven South Asian countries. The maximum temperature range was observed for Pakistan, followed by India, Bangladesh, Sri Lanka, Maldives, Bhutan and Nepal.

#### Inference

Hence, from the above graph we can infer that the average of almost all the South Asian Countries is on the higher side lying between 25°C to 40°C (approximately), thereby indicating towards a probable correlation between average temperature and the spread of Covid-19 infection.

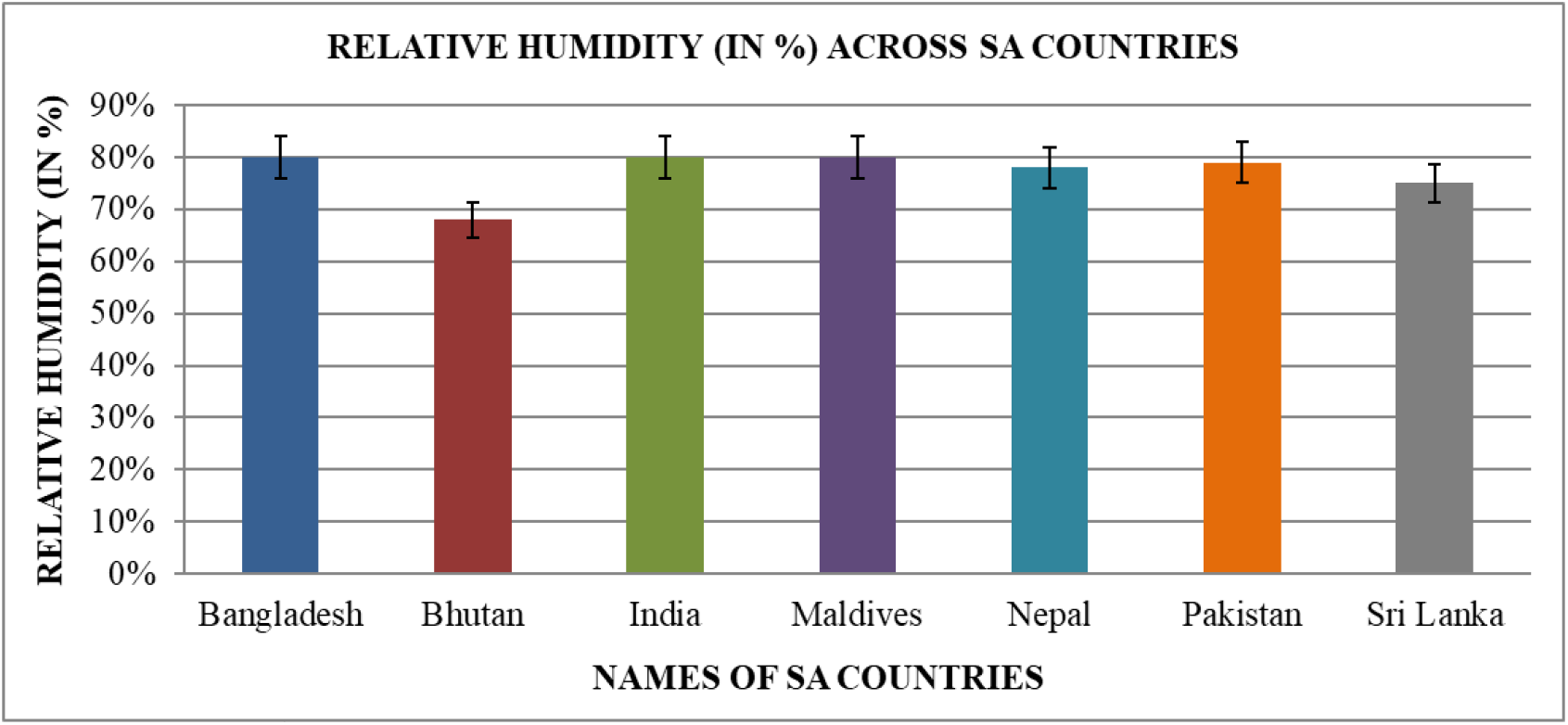

The above graph gives an account of the Relative humidity in each of the seven South Asian countries. The maximum relative Humidity was observed for Bangladesh, India and Maldives consecutively closely followed by Pakistan, Nepal, Sri Lanka and Bhutan.

#### Inference

Hence, from the above graph we can infer that the relative humidity of almost all the South Asian countries is on the higher side (within 65 to 80% approximately), with India, Maldives and Bangladesh recording the highest relative humidity around 80%, thereby indicating towards the possibility of a correlation between relative humidity and the spread of Covid-19 infection.

### 4.2. Data analysis using R Studio

#### 4.2.1. Country wise correlation of population density and Covid-19 related factors on 22^nd^ March

The Pearson’s correlation coefficient between the total number of cases and the population density per square kilometer of the different South Asian countries, was found out to be - 0.31455 which was statistically significant and indicated weak negative correlation. (95 percent confidence interval: −0.8971026 −0.6673755, t = −0.66275, df = 4, p-value = 0.5437)

The Pearson’s correlation coefficient between the total number of active cases and the population density per square kilometer of the different South Asian countries, was found out to be −0.31676 which was statistically significant and indicated weak negative correlation. (95 percent confidence interval: −0.8975793 −0.6660159, t = −0.6679, df = 4, p-value = 0.5408)

The Pearson’s correlation coefficient between the total number of deaths and the population density per square kilometer of the different South Asian countries, was found out to be −0.20963 which was statistically significant and indicated weak negative correlation. (95 percent confidence interval: −0.8727168 −0.7253320, t = −0.42878, df = 4, p-value = 0.6902)

The Pearson’s correlation coefficient between the total number of recoveries and the population density per square kilometer of the different South Asian countries, was found out to be −0.1626 which was statistically significant and indicated weak negative correlation. (95 percent confidence interval: −0.8605958 −0.7476188, t = −0.32958, df = 4, p-value = 0.7583)

#### 4.2.2. Country wise correlation of population density and Covid-19 related factors on 23^rd^ April

The Pearson’s correlation coefficient between the total number of cases and the population density per square kilometer of the different South Asian countries, was found out to be −0.2019057 which was statistically significant and indicated weak negative correlation. (95 percent confidence interval: −0.8707817 −0.7291300, t = −0.4123, df = 4, p-value = 0.7013)

The Pearson’s correlation coefficient between the total number of active cases and the population density per square kilometer of the different South Asian countries, was found out to be −0.1850397 which was statistically significant and indicated weak negative correlation. (95 percent confidence interval: −0.8664807 −0.7372326, t = −0.37658, df = 4, p-value = 0.7256)

The Pearson’s correlation coefficient between the total number of deaths and the population density per square kilometer of the different South Asian countries, was found out to be −0.1688403 which was statistically significant and indicated weak negative correlation. (95 percent confidence interval: −0.8622521 −0.7447730, t = −0.3426, df = 4, p-value = 0.7491)

The Pearson’s correlation coefficient between the total number of recoveries and the population density per square kilometer of the different South Asian countries, was found out to be −0.2593737 which was statistically significant and indicated weak negative correlation. (95 percent confidence interval: −0.8847064 −0.6994122, t = −0.53713, df = 4, p-value = 0.6197)

#### 4.2.2. Country wise correlation of population density and Covid-19 related factors on 25^nd^ May

The Pearson’s correlation coefficient between the total number of cases and the population density per square kilometer of the different South Asian countries, was found out to be −0.1506672 which was statistically significant and indicated weak negative correlation. (95 percent confidence interval: −0.8573905 −0.7529632, t = −0.30481, df = 4, p-value = 0.7757)

The Pearson’s correlation coefficient between the total number of active cases and the population density per square kilometer of the different South Asian countries, was found out to be −0.1246277 which was statistically significant and indicated weak negative correlation. (95 percent confidence interval: -0.8501975 - 0.7642301, t = -0.25121, df = 4, p-value = 0.814)

The Pearson’s correlation coefficient between the total number of deaths and the population density per square kilometer of the different South Asian countries, was found out to be - 0.1790794 which was statistically significant and indicated weak negative correlation. (95 percent confidence interval: -0.8649361 - 0.7400339, t = -0.36404, df = 4, p-value = 0.7343)

The Pearson’s correlation coefficient between the total number of recoveries and the population density per square kilometer of the different South Asian countries, was found out to be - 0.1801961 which was statistically significant and indicated weak negative correlation. (95 percent confidence interval: -0.8652265 - 0.7395115, t = -0.36639, df = 4, p-value = 0.7326)

#### 4.2.3. Country wise correlation of population density and Covid-19 related factors on 26^th^ June

The Pearson’s correlation coefficient between the total number of cases and the population density per square kilometer of the different South Asian countries, was found out to be - 0.1562078 which was statistically significant and indicated weak negative correlation. (95 percent confidence interval: -0.8588862 - 0.7504955, t = -0.3163, df = 4, p-value = 0.7676)

The Pearson’s correlation coefficient between the total number of active cases and the population density per square kilometer of the different South Asian countries, was found out to be -0.15198 which was statistically significant and indicated weak negative correlation. (95 percent confidence interval: -0.8577448 - 0.7523827, t = -0.30752, df = 4, p-value = 0.7738)

The Pearson’s correlation coefficient between the total number of deaths and the population density per square kilometer of the different South Asian countries, was found out to be - 0.1780369 which was statistically significant and indicated weak negative correlation. (95 percent confidence interval: -0.8646646 - 0.7405206, t = -0.36185, df = 4, p-value = 0.7358)

The Pearson’s correlation coefficient between the total number of recoveries and the population density per square kilometer of the different South Asian countries, was found out to be -0.1549712 which was statistically significant and indicated weak negative correlation. (95 percent confidence interval: -0.8585534 - 0.7510484, t = -0.31373, df = 4, p-value = 0.7694)

#### 4.2.4. Country wise correlation of population density and Covid-19 related factors on 27^th^ July

The Pearson’s correlation coefficient between the total number of cases and the population density per square kilometer of the different South Asian countries, was found out to be - 0.1425452 which was statistically significant and indicated weak negative correlation. (95 percent confidence interval: -0.8551762 - 0.7565352, t = -0.28803, df = 4, p-value = 0.7876)

The Pearson’s correlation coefficient between the total number of active cases and the population density per square kilometer of the different South Asian countries, was found out to be -0.08486247 which was statistically significant and indicated a very weak negative correlation. (95 percent confidence interval: -0.8386640 - 0.7804492, t = -0.17034, df = 4, p-value = 0.873)

The Pearson’s correlation coefficient between the total number of deaths and the population density per square kilometer of the different South Asian countries, was found out to be - 0.1629579 which was statistically significant and indicated weak negative correlation. (95 percent confidence interval: -0.8606923 - 0.7474545, t = -0.33033, df = 4, p-value = 0.7577)

The Pearson’s correlation coefficient between the total number of recoveries and the population density per square kilometer of the different South Asian countries, was found out to be -0.1709587 which was statistically significant and indicated weak negative correlation. (95 percent confidence interval: -0.8628106 - 0.7438000, t = -0.34703, df = 4, p-value = 0.7461)

#### 4.2.5. Time dependence of the Covid-19 related factors in Bangladesh

The Covid related data in Bangladesh like the total active cases, recovered cases and death cases were recorded at intervals of 15 days starting from 22^nd^ March, 2020 to 27^th^ July, 2020 and the Pearson’s correlation coefficient was determined taking two of the variables at a time.

The Pearson’s correlation coefficient between the total number of active cases and the total number of deaths of Bangladesh, was found out to be 0.972012 which was statistically significant and indicated a very strong positive correlation. (95 percent confidence interval: 0.8686037 - 0.9942872, t = 10.947, df = 7, p-value = 1.175e-05)

The Pearson’s correlation coefficient between the total number of active cases and time of Bangladesh, was found out to be 0.9378819 which was statistically significant and indicated a strong positive correlation. (95 percent confidence interval: 0.7258995 - 0.9871437, t = 7.1519, df = 7, p-value = 0.000185)

The Pearson’s correlation coefficient between the total number of deaths and time of Bangladesh, was found out to be 0.9411738 which was statistically significant and indicated a strong positive correlation. (95 percent confidence interval: 0.7389120 - 0.9878414, t = 7.3689, df = 7, p-value = 0.0001534)

The Pearson’s correlation coefficient between the total number of recovered cases and time of Bangladesh, was found out to be 0.8734471 which was statistically significant and indicated a strong positive correlation. (95 percent confidence interval: 0.4984831 - 0.9730985, t = 4.746, df = 7, p-value = 0.002093)

The Pearson’s correlation coefficient between the total number of recovered cases and active cases of Bangladesh, was found out to be 0.9095185 which was statistically significant and indicated a strong positive correlation. (95 percent confidence interval: 0.6197372 - 0.9810535, t = 5.7892, df = 7, p-value = 0.0006709)

The Pearson’s correlation coefficient between the total number of cases and time of Bangladesh, was found out to be 0.9378819 which was statistically significant and indicated a strong positive correlation. (95 percent confidence interval: 0.7258995 - 0.9871437, t = 7.1519, df = 7, p-value = 0.000185)

#### 4.2.6. Time dependence of the Covid-19 related factors in India

The Covid related data in India like the total active cases, recovered cases and death cases were recorded at intervals of 15 days starting from 22^nd^ March, 2020 to 27^th^ July, 2020 and the Pearson’s correlation coefficient was determined taking two of the variables at a time.

The Pearson’s correlation coefficient between the total number of active cases and the total number of deaths of India, was found out to be 0.9938517 which was statistically significant and indicated a very strong positive correlation. (95 percent confidence interval: 0.9699039 0.9987560, t = 23.749, df = 7, p-value = 5.965e-08)

The Pearson’s correlation coefficient between the total number of active cases and time of India, was found out to be 0.9120734 which was statistically significant and indicated a strong positive correlation. (95 percent confidence interval: 0.6288844 - 0.9816079, t = 5.8853, df = 7, p-value = 0.0006085)

The Pearson’s correlation coefficient between the total number of deaths and time of India, was found out to be 0.9137647 which was statistically significant and indicated a strong positive correlation. (95 percent confidence interval: 0.6349831 - 0.9819743, t = 5.9511, df = 7, p-value = 0.0005694)

The Pearson’s correlation coefficient between the total number of recovered cases and time of India, was found out to be 0.8552941 which was statistically significant and indicated a strong positive correlation. (95 percent confidence interval: 0.4425487 - 0.9690032, t = 4.3673, df = 7, p-value = 0.003285)

The Pearson’s correlation coefficient between the total number of recovered cases and active cases of India, was found out to be 0.9907427 which was statistically significant and indicated a strong positive correlation. (95 percent confidence interval: 0.9549586 - 0.9981246, t = 19.309, df = 7, p-value = 2.491e-07)

The Pearson’s correlation coefficient between the total number of cases and time of India, was found out to be 0.8772256 which was statistically significant and indicated a strong positive correlation. (95 percent confidence interval: 0.5105318 - 0.9739431, t = 4.8345, df = 7, p-value = 0.00189)

The Pearson’s correlation coefficient between the total number of active cases and new recovered of India, was found out to be 0.992865 which was statistically significant and indicated a strong positive correlation. (95 percent confidence interval: 0.9651414 - 0.9985558, t = 22.03, df = 7, p-value = 1.003e-07)

The Pearson’s correlation coefficient between the total number of active cases and new cases of India, was found out to be 0.9994253 which was statistically significant and indicated a strong positive correlation. (95 percent confidence interval: 0.9971558 0.9998840, t = 78.005, df = 7, p-value = 1.498e-11)

The Pearson’s correlation coefficient between the total number of new cases and time of India, was found out to be 0.9057422 which was statistically significant and indicated a strong positive correlation. (95 percent confidence interval: 0.6063602 0.9802319, t = 5.6541, df = 7, p-value = 0.0007712)

The Pearson’s correlation coefficient between the total number of new recovered cases and time of India, was found out to be 0.8601303 which was statistically significant and indicated a strong positive correlation. (95 percent confidence interval: 0.4571429 0.9701004, t = 4.4615, df = 7, p-value = 0.002931)

#### 4.2.7. Time dependence of the Covid-19 related factors in Pakistan

The Covid related data in Pakistan like the total active cases, recovered cases and death cases were recorded at intervals of 15 days starting from 22^nd^ March, 2020 to 27^th^ July, 2020 and the Pearson’s correlation coefficient was determined taking two of the variables at a time.

The Pearson’s correlation coefficient between the total number of active cases and the total number of deaths of Pakistan, was found out to be 0.6578803 which was statistically significant and indicated a moderate positive correlation. (95 percent confidence interval: -0.01108433 - 0.92002951, t = 2.3112, df = 7, p-value = 0.05409)

The Pearson’s correlation coefficient between the total number of active cases and time of Pakistan, was found out to be 0.7066771 which was statistically significant and indicated a positive correlation. (95 percent confidence interval: 0.08019034 - 0.93294803, t = 2.6425, df = 7, p-value = 0.0333)

The Pearson’s correlation coefficient between the total number of deaths and time of Pakistan, was found out to be 0.9516929 which was statistically significant and indicated a strong positive correlation. (95 percent confidence interval: 0.7815282 - 0.9900583, t = 8.2004, df = 7, p-value = 7.78e-05)

The Pearson’s correlation coefficient between the total number of recovered cases and time of Pakistan, was found out to be 0.8691133 which was statistically significant and indicated a strong positive correlation. (95 percent confidence interval: 0.4848396 - 0.9721265, t = 4.649, df = 7, p-value = 0.002345)

The Pearson’s correlation coefficient between the total number of recovered cases and active cases of Pakistan, was found out to be 0.3914553 which was statistically significant and indicated a weak positive correlation. (95 percent confidence interval: -0.3684552 - 0.8377768, t = 1.1255, df = 7, p-value = 0.2975)

The Pearson’s correlation coefficient between the total number of cases and time of Pakistan, was found out to be 0.9526069 which was statistically significant and indicated a strong positive correlation. (95 percent confidence interval: 0.7853078 - 0.9902500, t = 8.2851, df = 7, p-value = 7.284e-05)

The Pearson’s correlation coefficient between the total number of active cases and new recovered of Pakistan, was found out to be 0.6440306 which was statistically significant and indicated a moderate positive correlation. (95 percent confidence interval: -0.03510686 - 0.91625609, t = 2.2274, df = 7, p-value = 0.06121)

The Pearson’s correlation coefficient between the total number of active cases and new cases of Pakistan, was found out to be 0.7309035 which was statistically significant and indicated a moderate positive correlation. (95 percent confidence interval: 0.1297764 - 0.9391523, t = 2.8335, df = 7, p-value = 0.02528)

The Pearson’s correlation coefficient between the total number of new cases and time of Pakistan, was found out to be 0.5004199 which was statistically significant and indicated a moderate positive correlation. (95 percent confidence interval: -0.2451872 - 0.8740576, t = 1.5292, df = 7, p-value = 0.1701)

The Pearson’s correlation coefficient between the total number of new recovered cases and time of Pakistan, was found out to be 0.9159503 which was statistically significant and indicated a strong positive correlation. (95 percent confidence interval: 0.6429161 - 0.9824470, t = 6.0389, df = 7, p-value = 0.0005217)

#### 4.2.8. Time dependence of the Covid-19 related factors in Nepal

The Covid related data in Nepal like the total active cases, recovered cases and death cases were recorded at intervals of 15 days starting from 22^nd^ March, 2020 to 27^th^ July, 2020 and the Pearson’s correlation coefficient was determined taking two of the variables at a time.

The Pearson’s correlation coefficient between the total number of active cases and the total number of deaths of Nepal, was found out to be 0.8379122 which was statistically significant and indicated a strong positive correlation. (95 percent confidence interval: 0.3918397 - 0.9650224, t = 4.0617, df = 7, p-value = 0.004799)

The Pearson’s correlation coefficient between the total number of active cases and time of Nepal, was found out to be 0.8163038 which was statistically significant and indicated a strong positive correlation. (95 percent confidence interval: 0.3323661 - 0.9599906, t = 3.739, df = 7, p-value = 0.007272)

The Pearson’s correlation coefficient between the total number of deaths and time of Nepal, was found out to be 0.9178613 which was statistically significant and indicated a strong positive correlation. (95 percent confidence interval: 0.6499006 - 0.9828597, t = 6.1185, df = 7, p-value = 0.0004822)

The Pearson’s correlation coefficient between the total number of recovered cases and time of Nepal, was found out to be 0.799707 which was statistically significant and indicated a strong positive correlation. (95 percent confidence interval: 0.2891579 - 0.9560617, t = 3.5241, df = 7, p-value = 0.009674)

The Pearson’s correlation coefficient between the total number of recovered cases and active cases of Nepal, was found out to be 0.6094344 which was statistically significant and indicated a moderate positive correlation. (95 percent confidence interval: -0.09187107 - 0.90661430, t = 2.0337, df = 7, p-value = 0.08146)

The Pearson’s correlation coefficient between the total number of cases and time of Nepal, was found out to be 0.8970782 which was statistically significant and indicated a strong positive correlation. (95 percent confidence interval: 0.5762956 - 0.9783370, t = 5.3713, df = 7, p-value = 0.00104)

#### 4.2.9. Time dependence of the Covid-19 related factors in Bhutan

The Covid related data in Bhutan like the total active cases, recovered cases and death cases were recorded at intervals of 15 days starting from 22^nd^ March, 2020 to 27^th^ July, 2020 and the Pearson’s correlation coefficient was determined taking two of the variables at a time.

The Pearson’s correlation coefficient between the total number of active cases and the total number of deaths of Bhutan, could not be calculated due to insufficient data.

The Pearson’s correlation coefficient between the total number of active cases and time of Bhutan, was found out to be 0.4273153 which was statistically significant and indicated a moderate positive correlation. (95 percent confidence interval: -0.3306379 - 0.8501683, t = 1.2505, df = 7, p-value = 0.2513)

The Pearson’s correlation coefficient between the total number of deaths and time of Bhutan, could not be calculated due to insufficient data.

The Pearson’s correlation coefficient between the total number of recovered cases and time of Bhutan, was found out to be 0.8871034 which was statistically significant and indicated a fairly strong positive correlation. (95 percent confidence interval: 0.5427254 -0.9761384, t = 5.0849, df = 7, p-value = 0.001423)

The Pearson’s correlation coefficient between the total number of recovered cases and active cases of Bhutan, was found out to be 0.07297398 which was statistically significant and indicated a very weak positive correlation. (95 percent confidence interval: -0.6212560 - 0.7030246, t = 0.19359, df = 7, p-value = 0.852)

The Pearson’s correlation coefficient between the total number of cases and time of Bhutan, was found out to be 0.9586467 which was statistically significant and indicated a very strong positive correlation. (95 percent confidence interval: 0.8106001- 0.9915134, t = 8.912, df = 7, p-value = 4.547e-05)

#### 4.2.10. Time dependence of the Covid-19 related factors in Maldives

The Covid related data in Maldives like the total active cases, recovered cases and death cases were recorded at intervals of 15 days starting from 22^nd^ March, 2020 to 27^th^ July, 2020 and the Pearson’s correlation coefficient was determined taking two of the variables at a time.

The Pearson’s correlation coefficient between the total number of active cases and the total number of deaths of Maldives, was found out to be 0.4575434 which was statistically significant and indicated a moderate positive correlation. (95 percent confidence interval: - 0.2967502 - 0.8602620, t = 1.3614, df = 7, p-value = 0.2156)

The Pearson’s correlation coefficient between the total number of active cases and time of Maldives, was found out to be 0.5513275 which was statistically significant and indicated a moderate positive correlation. (95 percent confidence interval: -0.1779506 - 0.8896904, t = 1.7484, df = 7, p-value = 0.1239)

The Pearson’s correlation coefficient between the total number of deaths and time of Maldives, was found out to be 0.9644856 which was statistically significant and indicated a strong positive correlation. (95 percent confidence interval: 0.8355880 - 0.9927289, t = 9.6609, df = 7, p-value = 2.684e-05)

The Pearson’s correlation coefficient between the total number of recovered cases and time of Maldives, was found out to be 0.922293 which was statistically significant and indicated a strong positive correlation. (95 percent confidence interval: 0.6662741 - 0.9838141, t = 6.3136, df = 7, p-value = 0.0003989)

The Pearson’s correlation coefficient between the total number of recovered cases and active cases of Maldives, was found out to be 0.2347743 which was statistically significant and indicated a weak positive correlation. (95 percent confidence interval: -0.5086565 - 0.7776464, t = 0.63901, df = 7, p-value = 0.5431)

The Pearson’s correlation coefficient between the total number of cases and time of Maldives, was found out to be 0.9843447 which was statistically significant and indicated a very strong positive correlation. (95 percent confidence interval: 0.9247641 - 0.9968203, t = 14.776, df = 7, p-value = 1.557e-06)

#### 4.2.11. Correlation of country wise GDP with Covid-19 related factors

The effect of GDP per capita of the SA countries was taken into account to assess how efficiently the countries tackled the Covid situation. The correlation coefficient was determined between the GDP per capita and Covid related parameters like the total active cases, death cases, recovered cases, etc. and also the prevalent medical infrastructure.

The Pearson’s correlation coefficient between GDP per capita and percentage of the population tested was found out to be 0.7548569 which was statistically significant and indicated a moderate positive correlation. (95 percent confidence interval: 0.004168252 - 0.961403899, t = 2.5735, df = 5, p-value = 0.04983).

The Pearson’s correlation coefficient between GDP per capita and Help support available with respect to affected patients (beds/1000) was found out to be 0.8931319 which was statistically significant and indicated a strong positive correlation. (95 percent confidence interval: 0.4278109 -0.9842219, t = 4.44, df = 5, p-value = 0.006765).

The Pearson’s correlation coefficient between percentage below poverty line and mortality percentage was found out to be 0.6134337 which was statistically significant and indicated a moderate positive correlation. (95 percent confidence interval: -0.2595015 - 0.9347041, t = 1.7369, df = 5, p-value = 0.1429).

The Pearson’s correlation coefficient between GDP per capita and percentage recovered was found out to be 0.4659598 which was statistically significant and indicated a moderate positive correlation. (95 percent confidence interval: -0.4422986 - 0.9023781, t = 1.1776, df = 5, p-value = 0.292).

The Pearson’s correlation coefficient between Average Temperature and percentage of positive cases was found out to be 0.7894634 which was statistically significant and indicated a strong positive correlation. (95 percent confidence interval: 0.08978145 - 0.96739422, t = 2.876, df = 5, p-value = 0.03475).

The Pearson’s correlation coefficient between Humidity and percentage of positive cases was found out to be 0.5978859 which was statistically significant and indicated a moderate positive correlation. (95 percent confidence interval: -0.2822558 - 0.9315295, t = 1.6678, df = 5, p-value = 0.1562).

The Pearson’s correlation coefficient between GDP per capita and mortality percentage was found out to be -0.3903264 which was statistically significant and indicated a negative correlation. (95 percent confidence interval: -0.8836468 - 0.5137395, t = -0.94799, df = 5, p-value = 0.3867).

The Pearson’s correlation coefficient between percentage tested and percentage of positive cases was found out to be -0.3011048 which was statistically significant and indicated a weak negative correlation. (95 percent confidence interval: -0.8593140 - 0.5844849, t = -0.70606, df = 5, p-value = 0.5117).

The Pearson’s correlation coefficient between Average Temperature and mortality percentage was found out to be 0.8396544 which was statistically significant and indicated a strong positive correlation. (95 percent confidence interval: 0.2355136 - 0.9757423, t = 3.4569, df = 5, p-value = 0.0181).

The Pearson’s correlation coefficient between Help support available with respect to affected patients (beds/1000) and percentage of recovered was found out to be 0.6042968 which was statistically significant and indicated a moderate positive correlation. (95 percent confidence interval: -0.2729919 - 0.9328447, t = 1.6959, df = 5, p-value = 0.1507).

The Pearson’s correlation coefficient between Help support available with respect to affected patients (beds/1000) and mortality percentage was found out to be -0.4750148 which was statistically significant and indicated a moderate negative correlation. (95 percent confidence interval: -0.9045153 - 0.4328962, t = -1.207, df = 5, p-value = 0.2814).

The Pearson’s correlation coefficient between percentage below poverty line and percentage of positive cases was found out to be 0.5470857 which was statistically significant and indicated a moderate positive correlation. (95 percent confidence interval: -0.3502857 - 0.9207900, t = 1.4614, df = 5, p-value = 0.2037).

The Pearson’s correlation coefficient between GDP per capita and percentage of the population below poverty line was found out to be -0.6598167 which was statistically significant and indicated a moderate negative correlation. (95 percent confidence interval: -0.9438796 - 0.1853265, t = -1.9635, df = 5, p-value = 0.1068).

#### 4.2.12. The effect of population density on the pandemic

The effect of population density of the SA countries was taken into account to assess its impact on the transmission and severity of the virus. The correlation coefficient was determined between the population density and Covid related parameters like the total active cases, death cases, recovered cases, etc. and also the prevalent medical infrastructure.

The Pearson’s correlation coefficient between population density and the total number of cases was found out to be 0.9991003 which was statistically significant and indicated a strong positive correlation. (95 percent confidence interval: 0.9913842 - 0.9999064, t = 47.117, df = 4, p-value = 1.214e-06).

The Pearson’s correlation coefficient between population density and the total number of active cases was found out to be 0.9890723 which was statistically significant and indicated a strong positive correlation. (95 percent confidence interval: 0.8996678 - 0.9988577, t = 13.417, df = 4, p-value = 0.0001785).

The Pearson’s correlation coefficient between population density and the total number of death cases was found out to be 0.9991743 which was statistically significant and indicated a strong positive correlation. (95 percent confidence interval: 0.9920898 - 0.9999141, t = 49.184, df = 4, p-value = 1.023e-06).

The Pearson’s correlation coefficient between population density and the total number of recovered cases was found out to be 0.9952835 which was statistically significant and indicated a strong positive correlation. (95 percent confidence interval: 0.9555606 - 0.9995084, t = 20.519, df = 4, p-value = 3.332e-05).

## 5. Conclusion

In Bangladesh, from 22nd March 2020 to 27th July 2020, it was observed that the total number of cases, active cases, total number of deaths and recovered cases increased significantly with time (a very strong positive correlation). The recovered cases and the death cases were found to be positively related to the active cases.

In India, from 22nd March 2020 to 27th July 2020, it was observed that the total number of cases, active cases, total number of deaths and recovered cases increased significantly with time (a very strong positive correlation). The recovered cases and the death cases were found to be positively related to the active cases. It was also observed that with time the total number of new cases increased and the total number of new recovered was found to be positively correlated to both the total number of active cases and new discovered cases.

In Pakistan, from 22nd March 2020 to 27th July 2020, it was observed that the total number of cases, active cases, total number of deaths and recovered cases increased significantly with time (a very strong positive correlation). However, only a moderate positive correlation was obtained between the total number of active cases and total number of deaths, total number of active cases and total number of new recovered cases. With time, new recovered cases increased sharply although total number of new cases increased moderately.

In Nepal, from 22nd March 2020 to 27th July 2020, it was observed that the total number of cases, active cases, and total number of deaths increased significantly with time (a very strong positive correlation). The total of recovered cases was found to be moderately dependent on time and the active cases in Nepal whereas the total number of deaths was quite strongly dependent on the active cases.

In Bhutan, from 22nd March 2020 to 27th July 2020, the total number of cases and the total number of recovered cases increased substantially, although the number of active cases increased moderately over time. The total number of recovered cases was found to be very weakly though positively related to the active cases. Due to lack of data, we could not find any correlation between the number of deaths and active cases.

In Maldives, from 22nd March 2020 to 27th July 2020, it was observed that the total number of cases, recovered cases, and total number of deaths increased significantly with time (a strong positive correlation) and active cases increased moderately with time. The total number of deaths was moderately dependent on the number of active cases in Nepal whereas the recovered cases was weakly dependent.

A study of the GDP per capita of the South Asian countries showed that a higher GDP resulted in greater number of testing, a better medical and health support, a better recovery rate and a lower mortality rate. To assess the economic condition the percentage of population below poverty line was also taken into consideration and it was correlated with the above-mentioned criteria and it was observed that greater the proportion of financially disadvantage class more was their chance to contract the disease and succumb to it.

With the rise in average temperature of the SA countries, the percentage of positive cases was found to increase considerably whereas there was only a moderate positive correlation between the percentage of positive cases and humidity. It was also noted that percentage of mortality rose significantly with the rise in average temperature.

With a stronger Help support available with respect to affected patients (beds/1000), the percentage recovered increased moderately whereas the mortality percentage decreased. This indicates that an organized medical infrastructure with sufficient beds and amenities to treat Covid-19 affected patients was instrumental in reducing the mortality percentage.

The total number of active cases, death cases, recovered cases were strongly dependent on the population density of the SA countries.

## 6. Discussion

A consistent study of Covid-19 related factors like the total positive cases, death cases, active cases, population density, GDP, etc. for all the South Asian countries from 22nd March, 2020 to 27th July, 2020, hinted at a critical finding that the correlation between these factors were strongly dependent on time. So, initially, the population density played a major role in the spread of the disease. However, later, as considerable portion of the population got infected, the economic factors and medical infrastructure came to the forefront, critically determining the recovery and the mortality rates. The geographical location and topographical parameters are profound aspects that govern the contagious nature of the virus. For example, Nepal and Bhutan are Himalayan countries that are landlocked, restricting the modes of communication only to few buses and private transport (Tshokey et.al., 2020).

We took the GDP per capita of the South Asian countries into account to analyze how efficiently the government could deal with the pandemic situation in terms of providing better healthcare and medical support. It was seen that higher the GDP per capita better was the help support available with respect to affected patients (beds/1000). The number of people tested for Covid-19 was also positively dependent on the GDP per capita which meant that a stronger economy could allocate more funds to carry out a uniform testing across all economic and social stratas, more efficiently. A weak negative correlation was observed between percentage tested and the percentage of positive cases indicating that although initially the percentage tested positive increased with testing, gradually with a voluminous increase in the number of tests due to various interfering factors like panic and regular seasonal flu, it did not culminate in the compulsory increase of the number of Covid-19 positive cases. The number of recovered cases increased significantly with a rise in GDP across the countries whereas the number of deaths was indirectly proportional to the GDP. implying a stabler economy had better access to modern technology in the field of medicine to tackle the pandemic easily. The economic stability of a country is not just dependent on its GDP but is also a reflection of the number of people below the poverty line. The percentage of affected patients were observed to be moderately dependent on the number of people below the poverty line. It was seen that mortality was moderately dependent on the strength of the economically weaker section of the society which clearly indicated that the poor people had lesser access to the advanced medical infrastructure leading to their increased mortality.

The total number of active cases, death cases, recovered cases were strongly dependent on the population density of the SA countries. A higher population density particularly in the heavily crowded slum areas of the metropolitan cities showed a rapid spread of infection. Eg., the largest slum in the heart of Mumbai, India, Dharavi (Population density over 2,77,136/km2). Moreover, these are the places that harbor the economically poorer population who does not have an easy access to the health care facilities. So, a combined effect of these two forces that is cramming of a large number of people in a very small area and their substandard lifestyle drives them to higher risk of mortality.

We did this study based on limited data, so before strong establishment of the conclusion drawn as of now, more specific and detailed data followed by complete studies of further parameters are highly encouraged. Proper understanding of affecting parameters can focus on a new path for mitigating an epidemic like Covid-19.

## Data Availability

All the data referred to in the manuscript are available on request.

## Author’s approval

Authors have seen and approved the manuscript.

## Declaration

None.

## Reference

1. Abiad, A., Arao, R.M. and Dagli, S., 2020. The economic impact of the COVID-19 outbreak on developing Asia.

2. Afzal, I., Abdul Raheem, R., Rafeeq, N. and Moosa, S., 2020. Contact tracing for containment of novel coronavirus disease (COVID-19) in the early phase of the epidemic in the Maldives. Asia Pacific Journal of Public Health, p.1010539520956447.

3. Ahmad, T., Haroon, M.B. and Hui, J., 2020. Coronavirus Disease 2019 (COVID-19) Pandemic and economic impact. Pakistan journal of medical sciences, 36(COVID19- S4), p.S73.

4. Arfan, M., Shah, K., Abdeljawad, T., Mlaiki, N. and Ullah, A., 2020. A Caputo power law model predicting the spread of the COVID-19 outbreak in Pakistan. Alexandria Engineering Journal.

5. Barman MP, Rahman T, Bora K, Borgohain C. COVID-19 pandemic and its recovery time of patients in India: A pilot study. Diabetes Metab Syndr 2020;14:1205-11, doi: https://doi.org/10.1016/j.dsx.2020.07.004

6. Behnood, A., Golafshani, E.M. and Hosseini, S.M., 2020. Determinants of the infection rate of the COVID-19 in the US using ANFIS and virus optimization algorithm (VOA). Chaos, Solitons & Fractals, 139, p.110051.

7. Bhadra, A., Mukherjee, A. and Sarkar, K., 2020. Impact of population density on Covid-19 infected and mortality rate in India. Modeling Earth Systems and Environment, pp.1–7.

8. Case-Fatality Ratio and Recovery Rate of COVID-19: Scenario of Most Affected Countries and Indian States International Institute for Population Sciences, Mumbai (2020), doi: https://doi.org/10.1016/j.amsu.2020.06.041

9. Chalise, H.N., 2020. COVID-19 situation and challenges for Nepal. Asia Pacific Journal of Public Health, s32(5), pp.281–282.

10. Chalise, H.N., 2020. South Asia is more vulnerable to COVID-19 pandemic. Arch Psychiatr Ment Health, 4, p.046–047.

11. Coşkun, H., Yıldırım, N. and Gündüz, S., 2020. The spread of COVID-19 virus through population density and wind in Turkey cities. Science of the Total Environment, 751, pp.141663.

12. Fauci, A.S., Lane, H.C. and Redfield, R.R., 2020. Covid-19—navigating the uncharted.

13. Ferdous, M.Z., Islam, M.S., Sikder, M.T., Mosaddek, A.S.M., Zegarra-Valdivia, J.A. and Gozal, D., 2020. Knowledge, attitude, and practice regarding COVID-19 outbreak in Bangladesh: An online-based cross-sectional study. PloS one, 15(10), pp.e0239254.

14. Ghoushchi, S.J., Ahmadi, M., Sharifi, A., Dorosti, S., Jafarzadeh Ghoushchi, S. and Ghanbari, N., 2020. Investigation of effective climatology parameters on COVID-19 outbreak in Iran.

15. Gupta, A. and Pradhan, B., 2020. Impact of daily weather on COVID-19 outbreak in India. medRxiv.

16. Gupta, A., Pradhan, B. and Maulud, K.N.A., 2020. Estimating the impact of daily weather on the temporal pattern of COVID-19 outbreak in India. Earth Systems and Environment, 4(3), pp.523–534.

17. Haque, S.E. and Rahman, M., 2020. Association between temperature, humidity, and COVID-19 outbreaks in Bangladesh. Environmental science & policy, 114, p.253–255.

18. https://data.worldbank.org/indicator/NY.GDP.PCAP.CD?locations=8S

19. https://www.wikipedia.org/

20. Islam, S.D.U., Bodrud-Doza, M., Khan, R.M., Haque, M.A. and Mamun, M.A., 2020. Exploring COVID-19 stress and its factors in Bangladesh: A perception-based study. Heliyon, 6(7), p.e04399.

21. Joacim Rocklöv, PhD, Henrik Sjödin, PhD, High population densities catalyse the spread of COVID-19, Journal of Travel Medicine, Volume 27, Issue 3, April 2020, taaa038, https://doi.org/10.1093/jtm/taaa038

22. Johnson K. Census: 1 in 6 India city residents lives in slums. [Internet]. 22 Mar 2013. Available at:https://www.sandiegouniontribune.com/sdut-census-1-in-6-india-city-residents-lives-in-slums2013mar22-story.html. Accessed 21 Apr 2020.

23. Khafaie MA, Rahim F. Cross-Country Comparison of Case Fatality Rates of COVID- 19/SARS-COV-2. Osong Public Health Res Perspect. 2020;11(2):74-80. doi:10.24171/j.phrp.2020.11.2.03

24. Khan, S., Khan, M., Maqsood, K., Hussain, T. and Zeeshan, M., 2020. Is Pakistan prepared for the COVID-19 epidemic? A questionnaire-based survey. Journal of Medical Virology.

25. KM O’reilly, M Auzenbergs, Y Jafari, Y Liu, S Flasche, R Lowe Effective transmission across the globe: the role of climate in COVID-19 mitigation strategies Lancet Planet Health, 4 (2020), p. e172

26. Kodera, S., Rashed, E.A. and Hirata, A., 2020. Correlation between COVID-19 morbidity and mortality rates in Japan and local population density, temperature, and absolute humidity. International journal of environmental research and public health, 17(15), pp.5477.

27. Kopec, J.A., Finès, P., Manuel, D.G. et al.. Validation of population-based disease simulation models: a review of concepts and methods. BMC Public Health 10, 710 (2010). https://doi.org/10.1186/1471-2458-10-710

28. L Morawska, JW Tang, W Bahnfleth, et al. How can airborne transmission of COVID- 19 indoors be minimised? Environ Int, 142 (2020), Article 105832

29. Liang, L., Tseng, C., Ho, H.J. et al.. Covid-19 mortality is negatively associated with test number and government effectiveness. Sci Rep 10, 12567 (2020). https://doi.org/10.1038/s41598-020-68862-x

30. Liu et al., 2017 L. Liu, Y. Li, P.V. Nielsen, J. Wei, R.L. Jensen Short-range airborne transmission of expiratory droplets between two people Indoor Air, 27 (2017), pp. 452– 462

31. Maliszewska, M., Mattoo, A. and Van Der Mensbrugghe, D., 2020. The potential impact of COVID-19 on GDP and trade: A preliminary assessment.

32. Mckibbin, W.J. and Fernando, R., 2020. The global macroeconomic impacts of COVID-19: Seven scenarios.

33. Measuring the Economic Risk of COVID-19

34. Moosa, S. and Usman, S.K., 2020. Nowcasting the COVID-19 epidemic in the Maldives. The Maldives National Journal of Research, 8(1), pp.18–28.

35. News B. Coronavirus: How India’s Kerala state “flattened the curve.” 2020; Available from: https://www.bbc.com/news/world-asia-india-52283748

36. Nick J Beeching, Tom E Fletcher, Mike B J Beadsworth, Covid-19: testing times, BMJ 2020;369:m1403. doi: https://doi.org/10.1136/bmj.m1403

37. Our World in Data, Coronavirus (Covid-19) Testing, https://ourworldindata.org/coronavirus-testing

38. P. Dhillon, C.S. Sampurna Kundu, U. Ram, L.K. Dwivedi, S. Yadav, S. Unisapatel J. A. et al. Poverty, Inequality & COVID-19: The Forgotten Vulnerable. Public Health 183, 110–111 (2020).

39. Ph.D. Joel Hellewell, et al.

40. Rashed, E.A., Kodera, S., Gomez-Tames, J. and Hirata, A., 2020. Influence of absolute humidity, temperature and population density on COVID-19 spread and decay durations: Multi-prefecture study in Japan. International journal of environmental research and public health, 17(15), pp.5354.

41. Roshana, M., Kaldeen, M. and Banu, A.R., 2020. Impact of COVID-19 outbreak on Sri Lankan Economy. Journal of Critical Reviews, 7(14), pp.2124–2133.

42. Rothan, H.A. and Byrareddy, S.N., 2020. The epidemiology and pathogenesis of coronavirus disease (COVID-19) outbreak. Journal of autoimmunity, p.102433.

43. SAARC disaster management center, coronavirus disease (COVID-19) SAARC Region. http://www.covid19-sdmc.org/

44. Samaddar, A., Gadepalli, R., Nag, V.L. and Misra, S., 2020. The enigma of low COVID-19 fatality rate in India. Frontiers in Genetics, 11, p.854.

45. Santosh, K.C., 2020. COVID-19 prediction models and unexploited data. Journal of medical systems, 44(9), pp.1–4.

46. Shoaib, M. and Abdullah, F., 2020. Risk Reduction of COVID-19 Pandemic in Pakistan. Social work in public health, 35(7), pp.557–568.

47. Singh, D.R., Sunuwar, D.R., Adhikari, B., Szabo, S. and Padmadas, S.S., 2020. The perils of COVID-19 in Nepal: Implications for population health and nutritional status. Journal of global health, 10(1).

48. Spinelli A, Pellino G. COVID-19 pandemic: perspectives on an unfolding Crisis. British Journal of Surgery. 2020;107(7):785–787. doi:10.1002/bjs.11627.

49. Sun, Z., Zhang, H., Yang, Y., Wan, H. and Wang, Y., 2020. Impacts of geographic factors and population density on the COVID-19 spreading under the lockdown policies of China. Science of The Total Environment, 746, pp.141347.

50. Suzana1, M., Moosa, S., Rafeeg, F.N. and Usman, S.K., 2020. Early measures for prevention and containment of COVID-19 in the Maldives: A descriptive analysis. Journal of Health and Social Sciences, 5(2), pp.251–264.

51. To, T., Zhang, K., Maguire, B., Terebessy, E., Fong, I., Parikh, S. and Zhu, J., 2020. Correlation of ambient temperature and COVID-19 incidence in Canada. Science of the Total Environment, 750, pp.141484.

52. Tshokey, T., Choden, J., Dorjee, K., Pempa, P., Yangzom, P., Gyeltshen, W., Wangchuk, S., Dorji, T. and Wangmo, D., 2020. Limited Secondary Transmission of the Novel Coronavirus (SARS-CoV-2) by Asymptomatic and Mild COVID-19 Patients in Bhutan. The American Journal of Tropical Medicine and Hygiene, p.tpmd200672.

53. Velavan, T.P. and Meyer, C.G., 2020. The COVID-19 epidemic. Tropical medicine & international health, 25(3), pp.278.

54. Wang, J., Tang, K., Feng, K. and Lv, W., 2020. High temperature and high humidity reduce the transmission of COVID-19. Available at SSRN 3551767.

55. Waris, A., Khan, A.U., Ali, M., Ali, A. and Baset, A., 2020. COVID-19 outbreak: current scenario of Pakistan. New Microbes and New Infections, p.100681.

56. Wickramaarachchi, T., Perera, S. and Jayasinghe, S., 2020. COVID-19 epidemic in Sri Lanka: A mathematical and computational modelling approach to control. medRxiv.

57. World Health Organization. Coronavirus (COVID-19). https://who.sprinklr.com/. Accessed 19 Apr 2020.

58. World Health Organization. Laboratory testing strategy recommendations for COVID- 19. 22 Mar 2020. https://apps.who.int/iris/bitstream/handle/10665/331509/WHO-COVID-19-lab_testing-2020.1-eng.pdf

59. Worldometer, coronavirus, COVID-19 coronavirus pandemic. https://www.worldometers.info/coronavirus/?

60. Zietz and Tatonetti, 2020 M. Zietz, N.P. Tatonetti, Testing the association between blood type and COVID-19 infection, intubation, and death, medRxiv (2020)

